# Safety and immunogenicity of anti-SARS CoV-2 vaccine SOBERANA 02 in homologous or heterologous scheme

**DOI:** 10.1101/2021.11.14.21266309

**Authors:** Maria Eugenia-Toledo-Romani, Leslihana Verdecia-Sánchez, Meybis Rodriguez-González, Laura Rodríguez-Noda, Carmen Valenzuela-Silva, Beatriz Paredes-Moreno, Belinda Sanchez-Ramirez, Rocmira Perez-Nicado, Raul González-Mugica, Tays Hernández-Garcia, Gretchen Bergado-Baez, Franciscary Pi-Estopiñán, Otto Cruz-Sui, Anitza Fraga-Quintero, Majela García-Montero, Ariel Palenzuela-Diaz, Gerardo Baro-Roman, Ivis Mendosa-Hernández, Sonsire Fernandez-Castillo, Yanet Climent-Ruiz, Darielys Santana-Mederos, Ubel Ramírez Gonzalez, Yanelda García-Vega, Beatriz Perez-Massón, Guang-Wu-Chen, Tammy Boggiano-Ayo, Eduardo Ojito-Magaz, Daniel G. Rivera, Yury Valdés-Balbín, Dagmar García-Rivera, Vicente Vérez-Bencomo, SOBERANA Research Group

**Affiliations:** “Pedro Kourí” Tropical Medicine Institute. Av “Novia del Mediodía”, Kv 6 1/2, La Lisa, Habana, 11400, Cuba; Clinic #1. 21 St. and 190, La Lisa. Habana, Cuba; Finlay Vaccine Institute. 21st Ave. N^º^ 19810 between 198 and 200 Streets, Atabey, Playa, Havana, Cuba; Cybernetics, Mathematics and Physics Institute. 15^th^ Street #55, Vedado, Plaza de la Revolución, La Habana 10400, Cuba; Centre of Molecular Immunology. 15^th^ Ave. and 216 Street, Siboney, Playa, Havana, Cuba; National Civil Defense Research Laboratory. San José de las Lajas, Mayabeque, Cuba; Centre for Immunoassays. 134 St and 25, Cubanacán, Playa, La Habana, 11600 Cuba; National Clinical Trials Coordinating Center. 5th Ave and 62, Miramar, Playa, Havana, Cuba; Chengdu Olisynn Biotech. Co. Ltd., and State Key Laboratory of Biotherapy and Cancer Center, West China Hospital, Sichuan University, Chengdu 610041, People’s Republic of China; Laboratory of Synthetic and Biomolecular Chemistry, Faculty of Chemistry, University of Havana, Havana 10400, Cuba

## Abstract

**Background:** SOBERANA 02 is a COVID-19 conjugate vaccine candidate based on SARS-CoV-2 recombinant RBD conjugated to tetanus toxoid. SOBERANA Plus antigen is dimeric-RBD. Here we report safety, reactogenicity and immunogenicity from phase I and IIa clinical trials using two-doses SOBERANA 02 (homologous protocol) and three-doses (homologous) or heterologous (with SOBERANA Plus) protocols.

**Method:** We performed an open-label, monocentric, sequential and adaptive phase I for evaluating safety, reactogenicity and exploring immunogenicity of SOBERANA 02 in two formulations (15 and 25 μg) in 40 subjects, 19–59 years old. Phase IIa was open-label including 100 volunteers 19–80 years, receiving two doses of SOBERANA 02-25 μg. In both trials, half of volunteers received a third dose of SOBERANA 02, half received a heterologous dose of SOBERANA Plus-50 μg. Primary outcomes were safety and reactogenicity. The secondary outcome was vaccine immunogenicity evaluated by anti-RBD IgG ELISA, molecular neutralization test of RBD:hACE2 interaction, live-virus neutralization test and specific T-cells response.

**Results:** The most frequent AE was local pain, other AEs had frequencies ≤ 5%. No serious related AEs were reported. Phase IIa confirmed the safety results in 60–80 years subjects. In phase-I SOBERANA 02-25µg elicited higher immune response than SOBERANA 02-15 µg; in consequence, the higher dose progressed to phase IIa. Phase IIa results confirmed the immunogenicity of SOBERANA 02-25 μg even in 60–80 age range. Two doses of SOBERANA02-25 μg elicited an immune response similar to that of the Cuban Convalescent Serum Panel; it was higher after both the homologous and heterologous third doses; the heterologous scheme showing a higher immunological response.

**Conclusions:** SOBERANA 02 was safe and immunogenic in persons aged 19–80 years, eliciting neutralizing antibodies and specific T cell response. Highest immune responses were obtained in the heterologous three doses protocol. Trial registry: https://rpcec.sld.cu/trials/RPCEC00000340 and https://rpcec.sld.cu/trials/RPCEC00000347

## Introduction

Safe and effective vaccines are urgently needed to globally control the spread of COVID-19 [1; 2]. Novel vaccines based on mRNA and adenovirus-vector platforms [3; 4; 5, 6; 7; 8] and more traditional vaccines—as whole inactivated virus or protein subunit vaccines— [9; 10; 11; 12] have fulfilled the required efficacy threshold (≥ 50%) [2] and received emergency use authorizations; however, less than 5% of doses administered worldwide have gone to low-income countries [13, 14]. More than 100 COVID-19 vaccines are under clinical evaluation [15]; their success would be essential for reducing inequity in vaccine distribution worldwide. Among them, vaccines based on SARS-CoV-2 protein subunits have shown significant advantages concerning safety and conservation conditions, becoming more affordable for low- and middle-income countries. [16]

SOBERANA 02 is a protein subunit conjugate vaccine in which RBD is conjugated to tetanus toxoid (TT), produced by the Finlay Vaccine Institute (IFV) and the Centre for Molecular Immunology (CIM) in Havana. This is the only anti-SARS-CoV-2 conjugate vaccine in the clinical pipeline of WHO [15]; it is supported by a vast experience at IFV on carbohydrate-protein conjugate vaccines [17, 18]. By conjugating RBD to TT, both humoral and cellular immune responses are potentiated; the conjugate exposes multiple RBM (receptor binding motif) where neutralizing epitopes predominate [19]. In laboratory animals, RBD-TT elicited a robust neutralizing antibody response, a Th1-polarized T-cell response and immune memory [20].

SOBERANA 02 started phase I [21] (October 30^th^, 2020) and phase IIa [22] (December 17th, 2020) clinical trials for evaluating safety and immunogenicity in a two-doses scheme, followed by a third dose of SOBERANA 02 or SOBERANA Plus. SOBERANA Plus has been successfully evaluated as booster for COVID-19 convalescents [23, 24]; here it is evaluated for the first time as third dose in an heterologous immunization scheme.

## METHODS

### Products under evaluation

SOBERANA 02 and SOBERANA PLUS are suspensions for injection. Both are subunit vaccines based SARS-CoV-2 RBD, sequence Arg319-Phe541-(His)_6_ bearing a flexible C-terminal fragment that includes unpaired Cys538, produced in genetically modified CHO cells. In SOBERANA 02, RBD is conjugated to the carrier protein tetanus toxoid (TT); in SOBERANA Plus, RBD is dimerized (d-RBD) through a Cys538–Cys538 interchain disulfide bridge. SOBERANA 02 and SOBERANA Plus (Table I) are produced under GMP conditions at the Finlay Vaccine Institute (IFV) and the Centre for Molecular Immunology (CIM), in Havana, Cuba.

**Table 1.** Composition of vaccine candidates.

| Ingredient | Vaccine candidates |  |
| --- | --- | --- |
|  | SOBERANA 02 | SOBERANA Plus |
| Antigen | SARS-CoV-2 RBD conjugated to tetanus toxoid, 15 µg or 25 µg RBD per 20 µg tetanus toxoid | SARS-CoV-2 RBD dimer (d-RBD), 50 µg |
| Aluminium hydroxide (alum) | 0.5 mg | 1.25 mg |
| Sodium chloride | 4.25 mg | 4.25 mg |
| Disodium hydrogen phosphate | 0.03 mg | 0.03 mg |
| Sodium dihydrogen phosphate | 0.02 mg | 0.02 mg |
| Water for injection | 0.5 ml | 0.5 ml |

### Participants and study design

Eligible participants were healthy persons according to clinical and laboratory criteria, aged 19–59 years (phase I) or 19–80 years (phase IIa) of both sexes, recruited through public advertisement at community or professional environment close to the clinical site (Clinic #1, La Lisa Municipality in Havana). The health condition was assessed during the screening visit, based on medical records, physical examination, and clinical and microbiological laboratory tests. Key exclusion criteria were history of SARS CoV-2 infection, acute diseases, congenital or acquired immunodeficiencies, personal history of liver or kidney failure, immunological treatment in the last three months, allergy to ingredients in the formulation, pregnancy, puerperium or breastfeeding (Supplemental Material, Appendix A-1 y A-2).

Phase I clinical trial was open-label, monocentric, sequential and adaptive, for evaluating safety and reactogenicity and exploring immunogenicity of SOBERANA 02. Forty volunteers were enrolled, randomly and sequentially assigned to two groups of 20, for receiving 28 days apart two doses of 15 µg or 25 µg of SOBERANA 02. One group received the first dose of SOBERANA 02-15 µg; after the first interim analysis of safety on day 7, the second group received the first dose of SOBERANA 02-25 µg. On day 56, half of volunteers randomly assigned to each group received the third dose of SOBERANA 02 (homologous group, same dosage as first immunization) and half received SOBERANA Plus (50 µg of d-RBD/alumina, heterologous group) (Figure 1).

**Figure 1.**
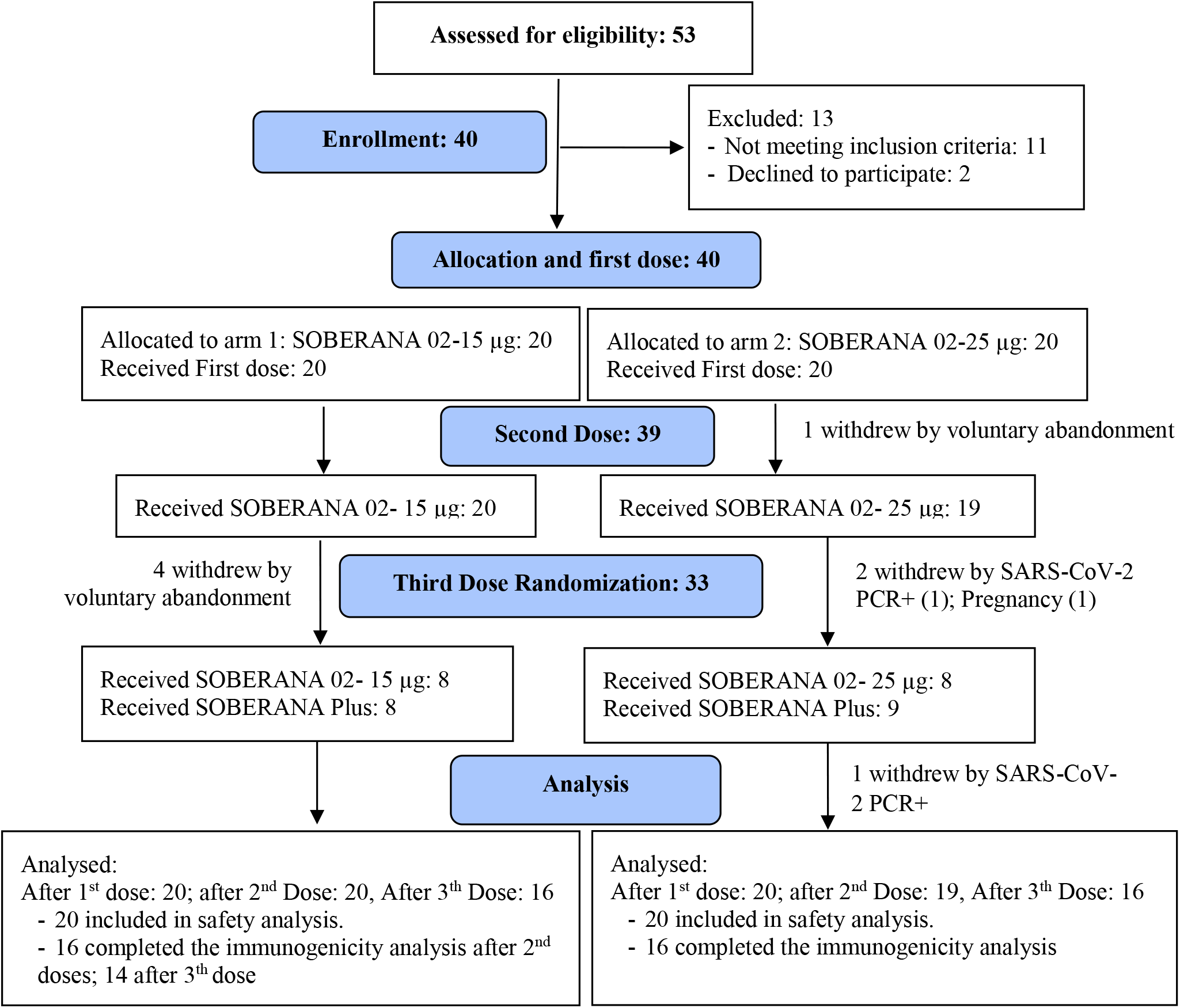
Phase I Flow Chart.

Phase II was an adaptive clinical trial for evaluating immunogenicity, safety and reactogenicity of SOBERANA 02. It was designed in two stages (IIa and IIb). Phase IIa started after the interim analysis (safety and preliminar immunogenicity) of phase I. It was open-label, including 100 volunteers aged 19–80 years (19–59: 76, 60–80: 24), receiving two doses of SOBERANA 02-25 µg on days 0 and 28. On day 56, participants were randomly allocated to receive either a third dose of SOBERANA 02-25 µg or SOBERANA Plus-50 µg (Figure 2). Phase IIb included 810 volunteers in a double blind, randomized, placebo controlled trial and will be published separately.

**Figure 2.**
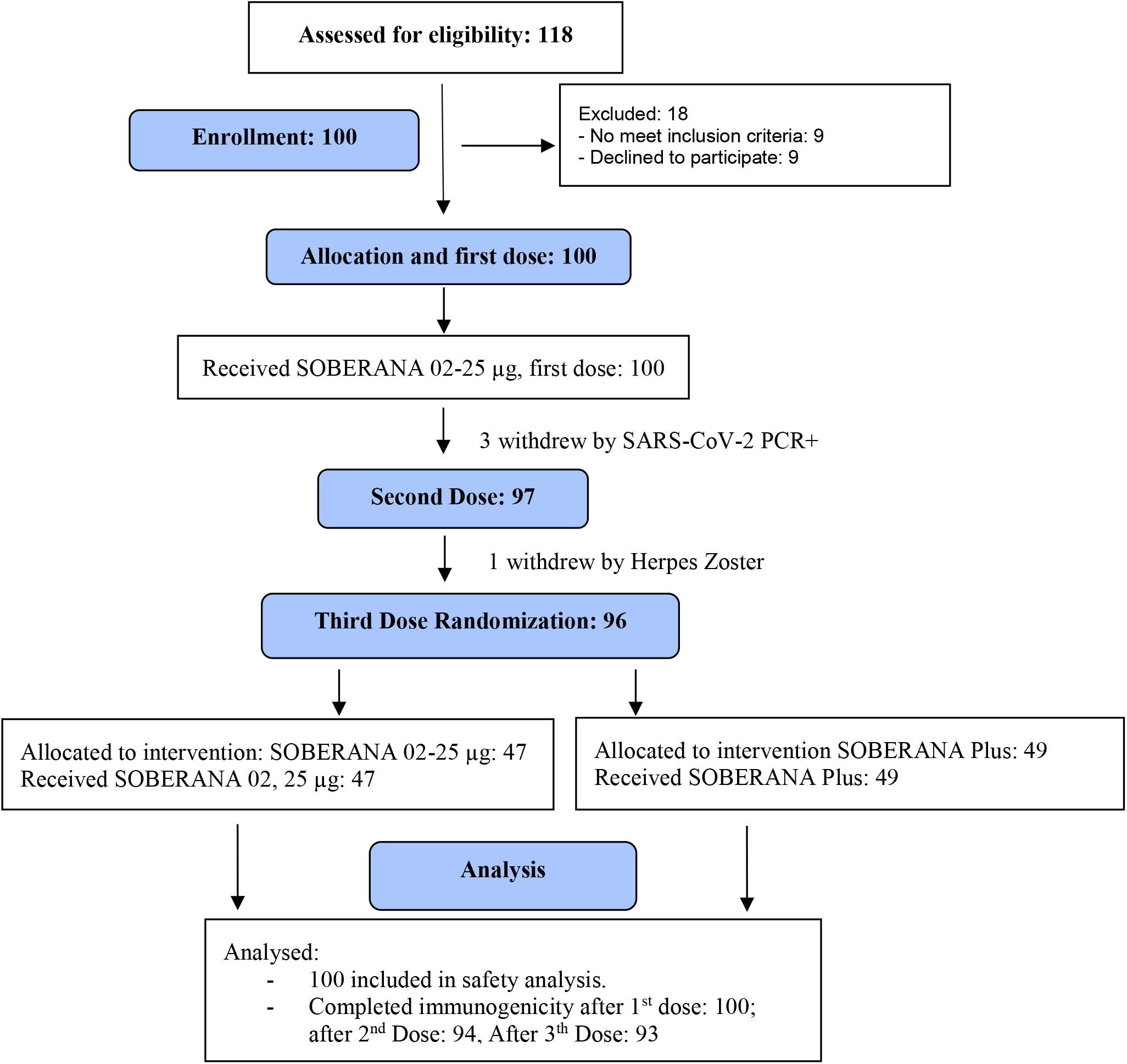
Phase IIa Flow Chart.

Both trials are published in the Cuban Public Registry of Clinical Trials, included in WHO International Clinical Registry Trials Platform with codes *<u>RPCEC00000340</u>* and *RPCEC00000347*. [21, 22]

### Ethical considerations

Phase I clinical trial was approved by the Ethical Committee at the Cuban National Centre for Toxicology; phase IIa was approved by a Research Ethic Committee from the Medical Sciences University, Faculty of Medicine “Manuel Fajardo”, Havana, designed by the Health Innovation Committee from the Cuban Ministry of Health (MINSAP). The Cuban National Regulatory Agency (Centre for State Control of Medicines and Medical Devices, CECMED) approved the trials and the procedures (CECMED, Authorizations dates: 29th October, 2020 for phase 1, Reference number: 05.014.20BA, and 17th December for phase 2, Reference number: 05.019.20BA).

Independent Data Monitoring Committees formed by external members (four in phase I and six in phase II committees) specialized on clinical practice, epidemiology and statistics were in charge of interim analysis of safety and immunogenicity. Three interim data analyses were done after first and second doses; satisfactory results allowed phase II trial authorization, incorporating elderly participants. The final data analysis was done 28 days post third immunization. The Cuban National Centre for the Coordination of Clinical Trials (CENCEC) was responsible for monitoring the trial in terms of adherence to the protocol, Good Clinical Practice and data accuracy.

Both trials were conducted according to Helsinki’s Declaration, Good Clinical Practice and the Cuban National Immunization Program. During recruitment, the investigators provided the potential participants with oral and written information about the vaccine candidates and trial potential risks and benefits. Written informed consent was obtained from all participants. The decision to participate was voluntary and was not remunerated.

### Procedures

Participants received intramuscular injections in the deltoid region. After each vaccination, they were closely followed for safety evaluation (during three hours in phase I and one hour in phase IIa). Medical visits were planned after each dose at 24, 48 and 72 hours, and on days 7 (in phase I), 14 and 28 (in phase I and IIa). Adverse event (AEs) were self-registered by the participants on a diary card and recorded during medical visits.

For evaluating immunogenicity, serum samples were collected on days 0 (before vaccination), 14, 28, 42, 56, 70 and 84 (this is, 14 and 28 days after each dose). Peripheral blood mononuclear cells were collected for T-cell response evaluation after the second dose (day 56) and after the third dose (day 84) in a participants’ subset.

### Outcomes

Both in phase I and phase IIa, the primary safety outcome was evaluated through the occurrence of serious AEs measured daily during 28 days after each dose. The secondary safety outcomes were solicited local and systemic AEs (measured daily during 7 days after each immunization) and unsolicited AE (measured daily during 28 days after each dose). Other secondary outcomes were vaccine immunogenicity, seroconversion (≥ 4-fold increase to pre-vaccination value), kinetics of anti-RBD IgG production (on days 0, 14, 28, 42, 56, 70 and 84), neutralizing antibody titres (on days 0, 56 and 84) and inhibition of RBD-ACE2 interaction (on days 0, 14, 28, 42, 56, 70 and 84). Outcomes are detailed in Supplemental Material, Appendix A-3)

### Safety evaluation

Solicited local AEs at the injection site included local pain, erythema, swelling, induration and local temperature; solicited systemic AEs were fever, general discomfort and rash. Other events were self-recorded throughout the 28 days follow-up period. Clinical laboratory test included pre-vaccination and post-vaccination biochemical serum analysis.

AEs were classified as serious or not. Also, AE severity was graded as mild (transient or mild discomfort, no interference with activity), moderate (mild to moderate limitation in activity), or severe (marked limitation in activity) according to Brighton Collaboration definition and the Common Terminology Criteria for Adverse Events version 5·0. AEs were reviewed for causality, and classified according to WHO: inconsistent causal association to immunization, consistent causal association to immunization, undetermined, unclassifiable [25].

### Immunogenicity evaluation

All immunological evaluations were performed by external laboratories.

#### Anti-RBD IgG response

Anti-RBD IgG in sera was evaluated by a quantitative ultramicro ELISA (UMELISA SARS-CoV-2 anti-RBD, Centre for Immunoassay, Havana, Cuba). The concentration of anti-RBD IgG was expressed as AU/mL. The seroconversion rate was calculated by dividing the concentration at each time point (at Tx) by the pre-vaccination concentration (at T0). A rate ≥ 4 was considered as seroconversion. (Supplemental Material, Appendix C.1)

#### Molecular virus neutralization test

This ELISA is an *in-vitro* surrogate of the live-virus neutralization with some modifications [26]. A molecular virus neutralization test with δ-variant L452R+T478K RBD displayed on phages was also evaluated. (Supplemental Material, Appendix C.2, C.3 and C.4) [27; 28]

#### Conventional virus neutralization test

Neutralizing antibodies against live D614G SARS-CoV-2 strain was performed by the conventional virus neutralization test, following the recommendation of Manenti & cols [29]. It is colorimetric assays based on the of the virus neutralization by antibodies, avoiding the cytopathic effect on VeroE6 cells. The neutralization titre represents the highest serum dilution giving 50% reduction of cytopathic effects. D614G strain was used for the test (Supplemental Material, Appendix C.5)

#### Specific T-cell response

RBD-specific T-cell response producing IFN-γ and IL-4 were quantified with enzyme-linked immunospot (ELISpot) assay using human IFN-γ ELISpot^PLUS^ HRP kit (Mabtech, Sweden) and human IL-4 ELISpot^plus^ HRP kit (Mabtech, Sweden) following the manufacturer’s instructions. Specific T-cell response was expressed as the number of spot-forming cells per 10^6^ cells. (Supplemental Material, Appendix C.6)

#### Human Serum Convalescent Panel

A panel of convalescent serum samples (Cuban Convalescent Serum Panel, CCSP) was made with sera from 68 patients recovered from COVID-19 (diagnosed by positive PCR) on March–November 2020, during the first epidemic peak in Cuba (13 with severe disease, 30 with mild disease and 25 asymptomatic). All patients gave written consent to the Cuban National Centre of Medical Genetics in Havana, allowing the use of their samples for epidemiological research. This panel was characterized by anti-RBD IgG concentration (UA/ml), inhibition of RBD-hACE2 interaction (% of inhibition and molecular neutralization titre) and virus neutralization titre (cVNT50) with the analytical methods used for vaccinated subjects in the clinical trials [23].

### Statistical analysis

Sample size calculation was done considering a serious AE rate < 5% (for phase I) and <1% (for phase IIa). Two-sided 95% confidence intervals for one proportion were calculated, taking into account a target width of 0.250 (for phase I) and 0.144 (for phase IIa). Safety and reactogenicity endpoints are described as frequencies (%). Demographic characteristics and AE data are reported as mean, standard deviation (SD), median, interquartile range, and range. Seroconversion rates for IgG antibodies anti-RBD (≥4-fold increase in antibody concentration over baseline) were calculated. Anti-RBD IgG concentration, inhibition (%) of RBD-ACE2 interaction and cytokine-expressing cells were represented as median with interquartile range. Molecular neutralization titre (mVNT_50_) and conventional virus neutralization titre (cVNT_50_) are represented as geometric mean (GMT) and 95% confidence intervals (CI). Spearman’s rank correlation was used to assess relationships among techniques used to evaluate the immune response. The Students’s t-Test or the Wilcoxon Signed-Rank Test were used for before-after statistical comparison. Statistical analyses were done using SPSS version 25·0; R version 3·2·4; EPIDAT version 4·1 and Prism GraphPad version 6·0. An alpha signification level of 0·05 was used. An Independent Data and Safety Monitoring Board provided safety supervision and interim analysis.

## Results

Phase I: from 53 individuals recruited for inclusion and screened from November 2^th^ to 12^th^, 2020, 40 participants were selected (Figure 1). The safety interim analysis for the group receiving SOBERANA 02-15 μg showed no serious AEs; then, the second group received SOBERANA 02-25 μg. Other two interim analyses (7 days after the second dose in the 15 μg-group and 7 days after the first dose in the 25 μg-group) showed no serious AEs. On day 56 half of individuals received a third dose of SOBERANA 02 (same dosage), half received a heterologous third dose of SOBERANA Plus-50 µg. Phase IIa: from 118 individuals recruited for inclusion and screened from December 17^th^ 2020 to January 6^th^, 2021, (Figure 2); the 100 selected participants received two doses of SOBERANA 02-25 μg. Demographic characteristics are summarized in Table 2. The mean age of participants was 38.2 years (SD 10.3) in phase I and 46.7 (SD 15.8) in phase IIa.

**Table 2.**
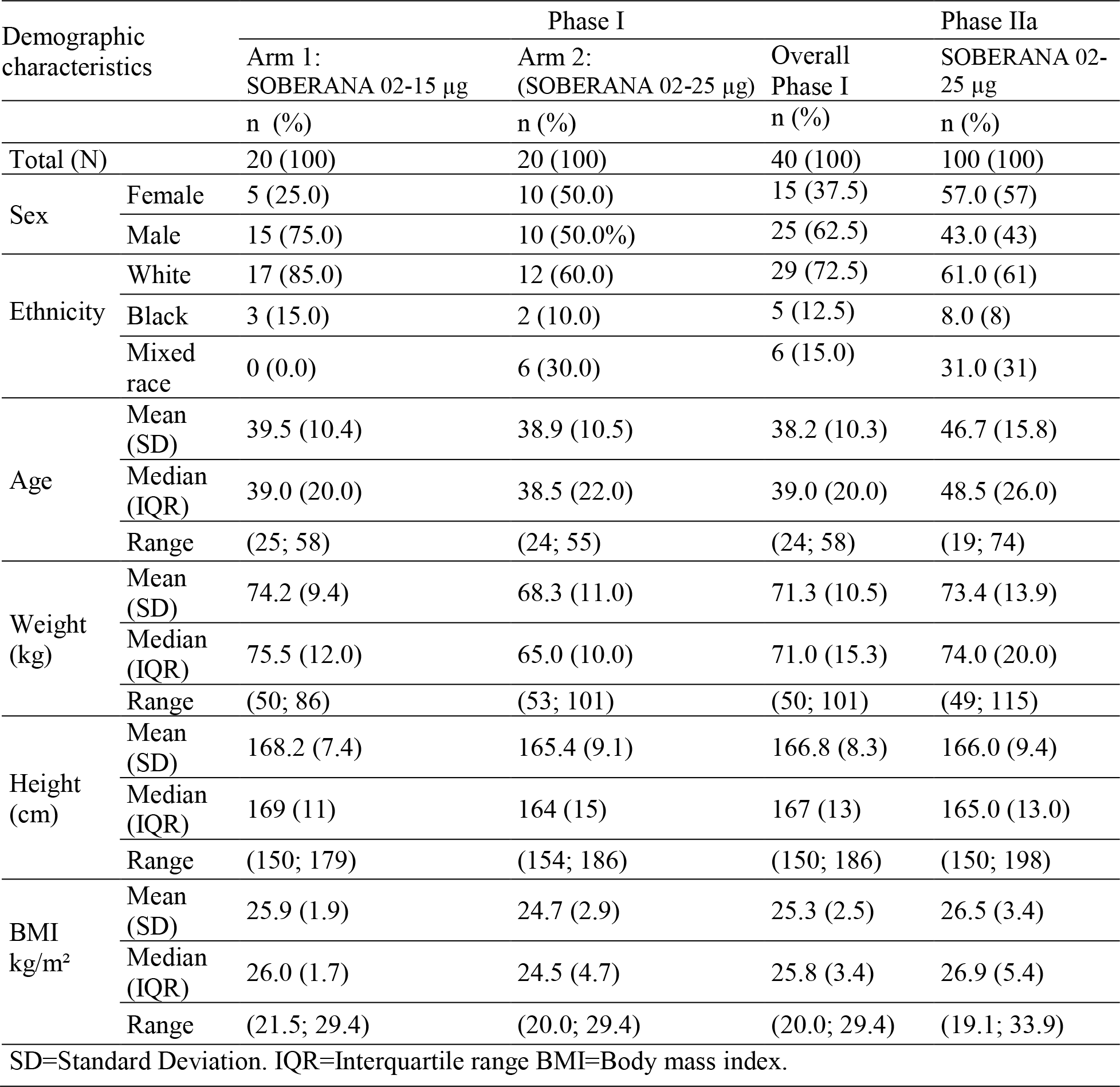
Demographic characteristics of participants in phase I and phase IIa clinical trials.

### Adverse events

Thirty of 40 participants in phase I (80%) and 93 of 100 in phase IIa completed the three-dose scheme and follow-up visits. In phase I, 16 (40%) reported at least one AE within 28 days after vaccination. In the group receiving SOBERANA 02-25 μg, 50% of subjects reported AEs compared to 30% in the group receiving SOBERANA 02-15 μg; none reported serious or severe (grade 3) vaccine-related AEs. In phase IIa, 32 participants (32%) reported at least one AE within 28 days after vaccination; none reported serious vaccine-related AEs and only one reported two severe (grade 3) AEs (induration and erythema, Tables 3 and 4). No clinically relevant changes were observed in haematology and blood chemistry analyses (Supplemental Material, Appendix B, Table IV).

**Table 3.**
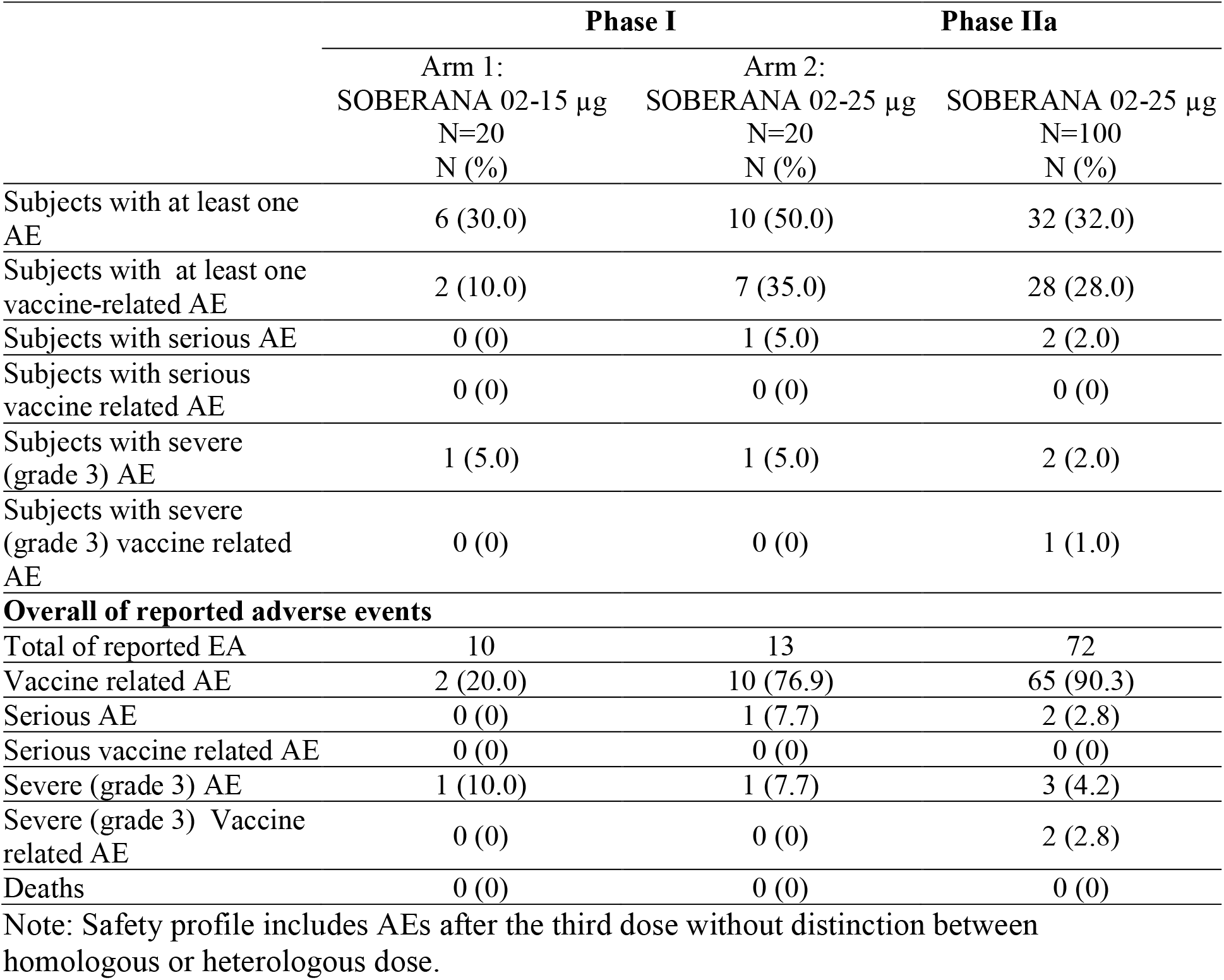
Phase I and phase IIa safety profile.

|  | Phase I |  | Phase IIa |
| --- | --- | --- | --- |
|  | Arm 1:<br>SOBERANA 02-15 µg<br>N=20<br>N (%) | Arm 2:<br>SOBERANA 02-25 µg<br>N=20<br>N (%) | SOBERANA 02-25 µg<br>N=100<br>N (%) |
| Subjects with at least one AE | 6 (30.0) | 10 (50.0) | 32 (32.0) |
| Subjects with at least one vaccine-related AE | 2 (10.0) | 7 (35.0) | 28 (28.0) |
| Subjects with serious AE | 0 (0) | 1 (5.0) | 2 (2.0) |
| Subjects with serious vaccine related AE | 0 (0) | 0 (0) | 0 (0) |
| Subjects with severe (grade 3) AE | 1 (5.0) | 1 (5.0) | 2 (2.0) |
| Subjects with severe (grade 3) vaccine related AE | 0 (0) | 0 (0) | 1 (1.0) |
| **Overall of reported adverse events** |  |  |  |
| Total of reported EA | 10 | 13 | 72 |
| Vaccine related AE | 2 (20.0) | 10 (76.9) | 65 (90.3) |
| Serious AE | 0 (0) | 1 (7.7) | 2 (2.8) |
| Serious vaccine related AE | 0 (0) | 0 (0) | 0 (0) |
| Severe (grade 3) AE | 1 (10.0) | 1 (7.7) | 3 (4.2) |
| Severe (grade 3) Vaccine related AE | 0 (0) | 0 (0) | 2 (2.8) |
| Deaths | 0 (0) | 0 (0) | 0 (0) |

Note: Safety profile includes AEs after the third dose without distinction between homologous or heterologous dose.

**Table 4.** Solicited AEs during phase I and phase IIa.

|  | **Phase I** |  | **Phase IIa** |
| --- | --- | --- | --- |
|  | Arm 1:<br>SOBERANA 02-15 µg<br>N=20<br>N (%) | Arm 2:<br>SOBERANA 02-25 µg<br>N=20<br>N (%) | SOBERANA 02-25 µg<br>N=100<br>N (%) |
| **Overall AE** |  |  |  |
| Subject with AEs | 6 (30.0) | 10 (50.0) | 32 (32.0) |
| Severe (Grade 3) | 1 (5.0) | 1 (5.0) | 2 (2.0) |
| Serious | 0 | 1 (5.0) | 2 (2.0) |
| **Subjects with solicited AEs** |  |  |  |
| Any | 0 | 3 (15.0) | 25 (25.0) |
| Severe (Grade 3) | 0 | 0 | 1 (1.0) |
| Serious | 0 | 0 | 0 |
| **Subjects with solicited systemic AEs** |  |  |  |
| Any | 0 | 0 | 6 (6) |
| Severe (Grade 3) | 0 | 0 | 0 |
| Serious | 0 | 0 | 0 |
| General discomfort | 0 | 0 | 5 (5) |
| Severe (Grade 3) | 0 | 0 | 0 |
| Serious | 0 | 0 | 0 |
| Rash | 0 | 0 | 1 (1) |
| Severe (Grade 3) | 0 | 0 | 0 |
| Serious | 0 | 0 | 0 |
| **Subjects with solicited local AEs** |  |  |  |
| Any | 0 | 3 (15) | 22 (22) |
| Severe (Grade 3) | 0 | 0 | 1 (1) |
| Serious | 0 | 0 | 0 |
| Injection-site pain | 0 | 3 (15) | 22 (22) |
| Severe (Grade 3) | 0 | 0 | 0 |
| Serious | 0 | 0 | 0 |
| Erythema | 0 | 0 | 4 (4) |
| Severe (Grade 3) | 0 | 0 | 1 (1) |
| Serious | 0 | 0 | 0 |
| Local Warm | 0 | 0 | 4 (4) |
| Severe (Grade 3) | 0 | 0 | 0 |
| Serious | 0 | 0 | 0 |
| Induration | 0 | 0 | 3 (3) |
| Severe (Grade 3) | 0 | 0 | 1 (1) |
| Serious | 0 | 0 | 0 |
| Swelling | 0 | 0 | 3 (3) |
| Severe (Grade 3) | 0 | 0 | 0 |
| Serious | 0 | 0 | 0 |

Table 3 summarizes the frequency of subjects with solicited AEs. In phase I, local pain was reported in three subjects receiving SOBERANA 02-25 μg (15%). Other events were systemic and unsolicited. The most frequent unsolicited AE in both treatment groups was high blood pressure (15% and 25% respectively) (Supplemental Material, Appendix B, Table I); of all AEs 70% (arm 1: 15 µg) and 84.6% (arm 2: 25 µg) were classified as mild (Supplemental Material, Appendix B, Table II). In Phase IIa, pain at the injection site was also the most frequent solicited AE (in 22% of subjects). Other solicited and unsolicited AEs had frequencies ≤ 5%. Headache was the most frequent vaccine-related, unsolicited AE (SM, Appendix B, Table II); of all AEs, 90.3% were classified as mild and 77.8% lasted < 24 hours. No serious related adverse event or death were reported during phases I and IIa. (Supplemental Material, Appendix B, Table III). The number of participants reporting AEs decreased with the number of doses. AEs behaved similarly in both age subgroups (19-59 and 60-80 years) (Figure 3).

**Figure 3.**
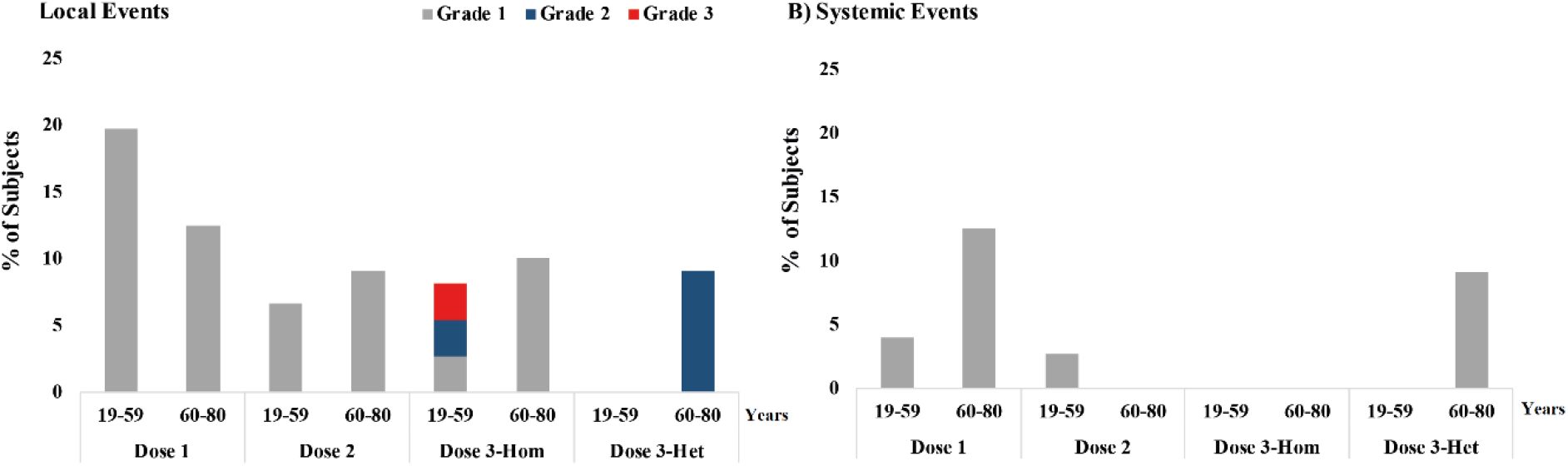
Solicited local and systemic adverse event after each dose by age subgroups. Subjects 19-59 or 60-80 years-old received two doses (T0, T28) of SOBERANA 02 and a third dose (T56) homologous (Hom) or heterologous with SOBERANA Plus (Het).

### Immunogenicity

In phase I, after the second dose both formulations of SOBERANA 02 induced seroconversion in ≥75% participants. The third dose increased seroconversion: 85.7% after the homologous third dose, 100% after the heterologous third dose (SOBERANA Plus) (Supplemental Material, Appendix B, Table V).

After two doses, the median of anti-RBD IgG concentration in subjects vaccinated with SOBERANA 02-15 μg was 25.9 (25^th^-75^th^ percentile 14.9; 39.5); in those vaccinated with SOBERANA 02-25 μg the median was 40.3 (25^th^-75^th^ percentile 18.5; 102.9) (Supplemental Material, Appendix B, Table V). Molecular inhibition of RBD:hACE2 interaction (expressed as % inhibition and molecular virus inhibitory titre 50%) was higher in the 25 μg-than in the 15 µg-group. Virus neutralization titre was 5.8 (95% CI 4.5; 7.5) after two doses of 15 μg, it was 21.7 (95% CI 7.8; 60.3) after two doses of 25 μg (Supplemental Material, Appendix B, Table V).

In all participants, the third dose increased the IgG concentration (p<0.05) as compared with the second dose. The combination of two doses of SOBERANA 02-25 μg with the heterologous third dose (SOBERANA Plus) also improved antibody functionality as compared with the homologous scheme: median of % inhibition of RBD:hACE2 interaction increased from 60.9% (25^th^-75^th^ percentile 11.9; 87.6) to 89.2% (25^th^-75^th^ percentile 57.2; 94.2), the GMT of molecular virus-neutralization titre (mVNT50) increased from 94.5 (95% IC 18.5; 481.2) to 340 (95% IC 125.8; 918.5) and the conventional live-virus neutralization increased form 24.2 (95% IC 9; 65.3) to 65.6 (95% IC 22; 195.8) (Supplemental Material, Appendix B, Table V).

Given the interim phase I results, phase IIa participants received SOBERANA 02-25µg in first and second immunizations, followed by homologous or heterologous third immunization. The study included participants up to 80 years in both schemes. The results were quite similar to those from phase I: 75% of participants seroconverted after the second dose and ≥ 95% after the third, with significant increment (p<0,05) in anti-RBD IgG titre, higher % inhibition of RBD:hACE2 interaction, higher molecular and virus neutralization titres, and better immunological results for the heterologous as compared with the homologous scheme (Supplemental Material, Appendix B, Table V).

The following are pooled data from all participants (in phases I and IIa) treated under the same vaccination scheme, two doses of SOBERANA 02-25 μg followed by either the homologous or the heterologous third dose. The proportion of participants that seroconverted increased from 76.1% after two doses (day 56) to 98.3% or 98% respectively after the third heterologous or homologous dose (day 84) (Supplemental Material, Appendix B, Table VI). A significant increase (p<0,0005) of anti-RBD antibodies was observed after first (day 14) and second doses (day 42) as compared with pre-vaccination (Figure 4). For both third dose subgroups, on day 84 the IgG level was significantly superior (p<0,0005) to its value on day 56; the highest increase was observed in subjects with the heterologous third dose (on day 84, the median with heterologous scheme was 4.7-fold higher than on day 56; whilst with the homologous scheme the increase was 3.4-fold). Also, after the heterologous third dose, the median IgG value was 2.2-fold higher than the median for the Cuban Convalescent Serum Panel (CCSP) (1.6-fold higher after the homologous third dose) (Figure 4; Supplemental Material, Appendix B, Table VI).

**Figure 4.**
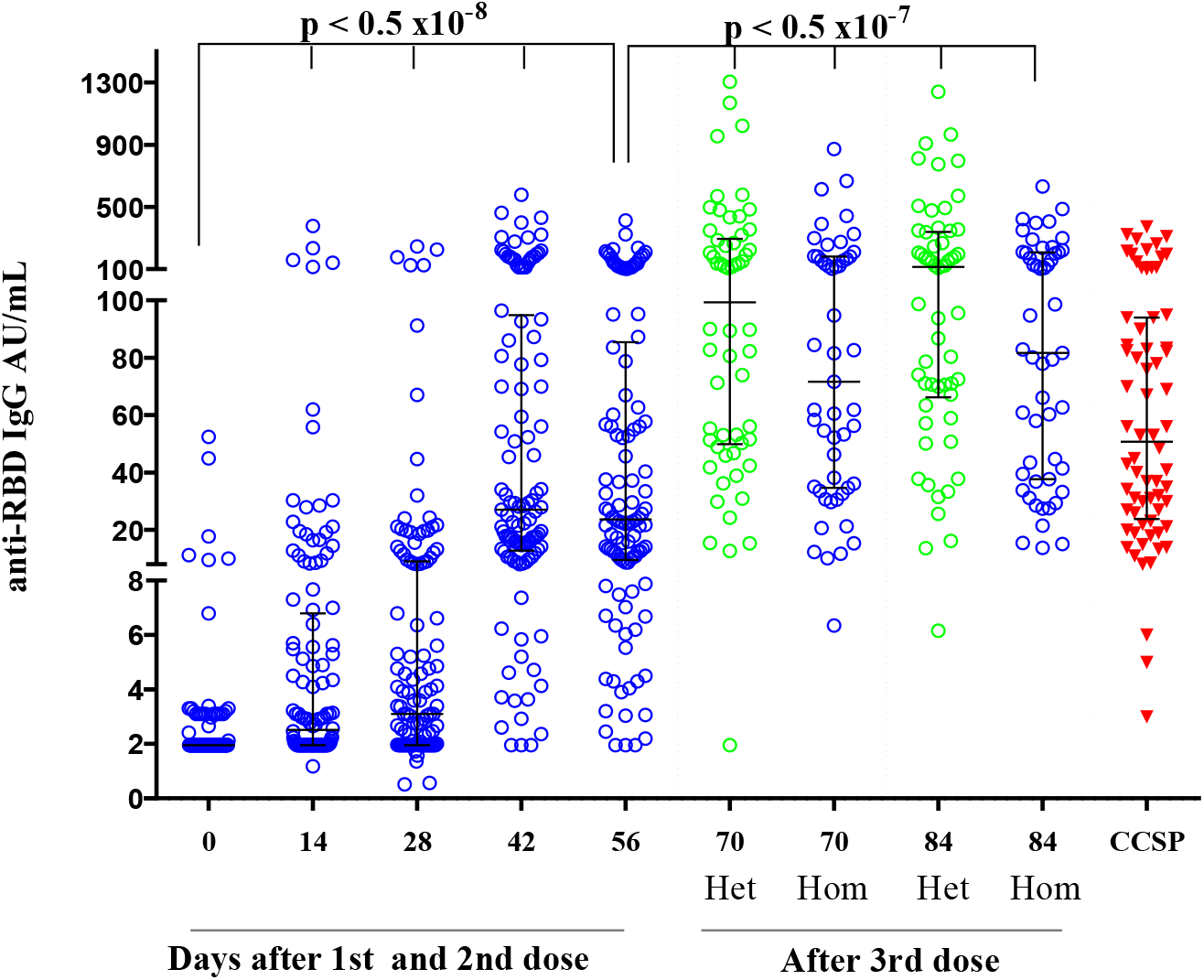
Kinetics of anti-RBD IgG production after two doses of SOBERANA 02 and a third homologous or heterologous dose (pooled analysis from phase I and phase IIa). FOOTNOTE: Subjects 19-80 years-old received two doses (T0, T28) of SOBERANA 02 and a third dose (T56) homologous (Hom: blue points) or heterologous with SOBERANA Plus (Het: green points). Anti-RBD IgG concentration is expressed in arbitrary units/mL (median, 25^th^-75^th^ percentile). CCSP: Cuban Convalescent Serum Panel (red triangles). p values represent the statistic differences respect to T0 or T56 as indicated, using Wilcoxon signed rank test.

Elicited anti-RBD antibodies inhibited the interaction of RBD with the human ACE2 receptor. There was a significant increase in % inhibition (p<0,0005) after the second dose (T42) compared to pre-vaccination and after both alternative third doses compared to T56 (Figure 5A). After the third dose (T84), the inhibition median was 78.9% (25^th^-75^th^ percentile 53.6; 91.1) and GMT of molecular neutralization titre was 257.7 (95% IC 203.2; 326.9), both significantly higher (p<0,0005) than those attained after the second dose: the heterologous third dose showed an mVNT_50_ increase of 5.7-fold, the homologous scheme increased 3.6-fold (Figure 5A, 5B; Supplemental Material, Appendix B, Table VII, Table VII). As observed in Figure 5B, GMT of mVNT50 after two doses was similar to the value for CCSP, and it was higher after the third dose, particularly after the heterologous immunization.

**Figure 5.**
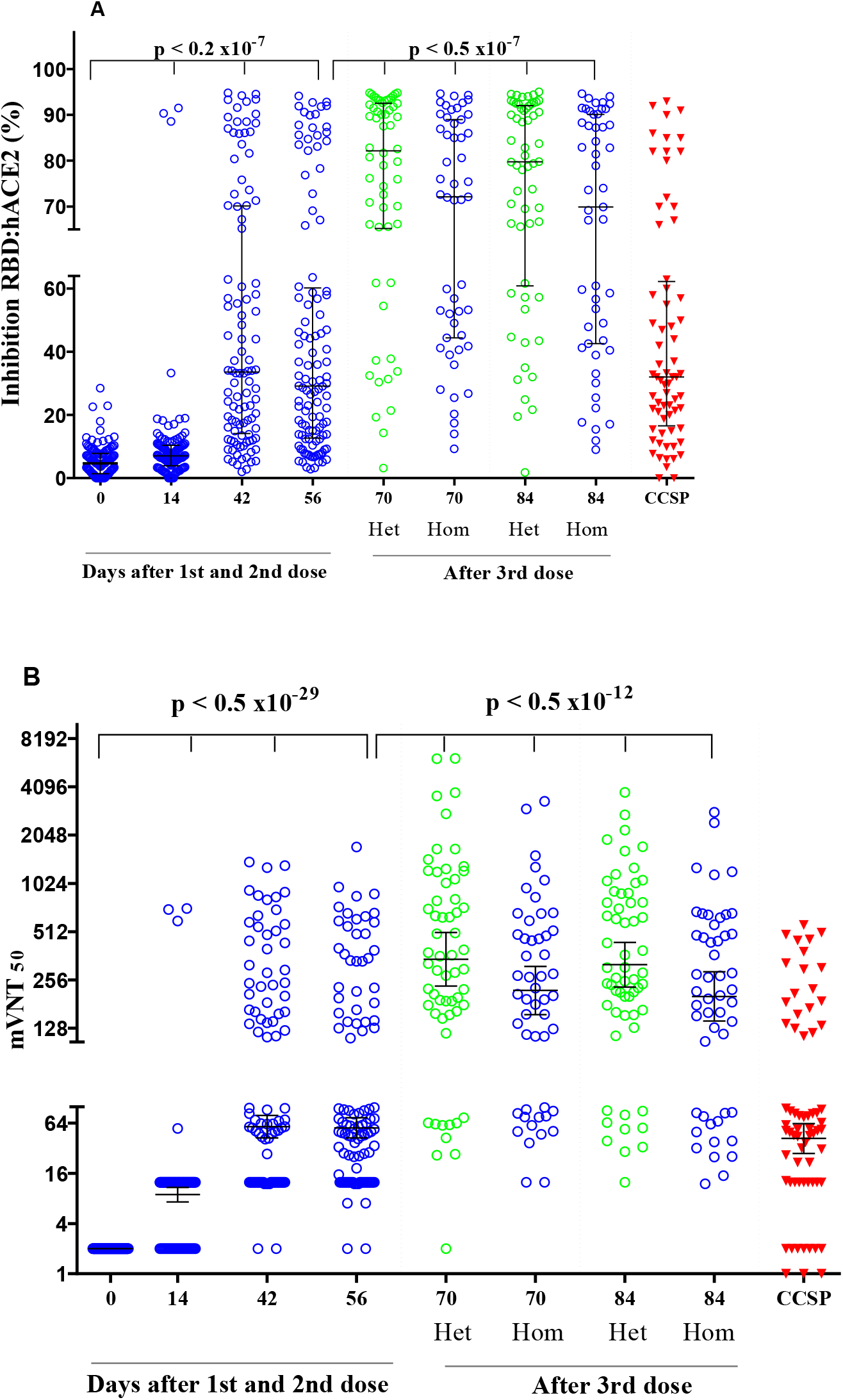
Anti-RBD IgG antibodies inhibit the interaction between RBD and human ACE2 receptor after two doses of SOBERANA 02 and a third homologous or heterologous dose (pooled analysis from phase I and phase IIa). **FOOTNOTE:** Subjects 19-80 years old received two doses (T0, T28) of SOBERANA 02 and a third dose (T56) homologous, (Hom: blue points) or heterologous with SOBERANA Plus (Het: green points). A: % inhibition of RBD:hACE2 interaction at 1/100 serum dilution (median, 25^th^-75^th^ percentile). B: Molecular virus neutralization titre mVNT50: highest serum dilution inhibiting 50% of RBD:hACE2 interaction; (GMT, IC 95%). CCSP: Cuban Convalescent Serum Panel (red triangles). p values represent statistic differences with T0 or T56, as indicated.

The conventional virus neutralization titre (cVNT_50_) was evaluated pre-vaccination and 28 days after the second and third doses (Figure 6). After two doses, the GMT reached 12.5 (95% IC 9.6; 16.1), significantly increasing (p<0,0005) to 37.5 (95% IC 29.8; 47.3) after the third dose. There were no significant differences for GMT cVNT_50_(heterologous: 42.5, 95% IC 30.4; 59.4), homologous: 32.8, 95% IC 23.8; 45.3); the heterologous third dose showed a cVNT_50_ increase of 3.4-fold, the homologous scheme increased 2.6-fold. They were similar to the CCSP value (Figure 6) (SM, Appendix B, Table VIII).

**Figure 6.**
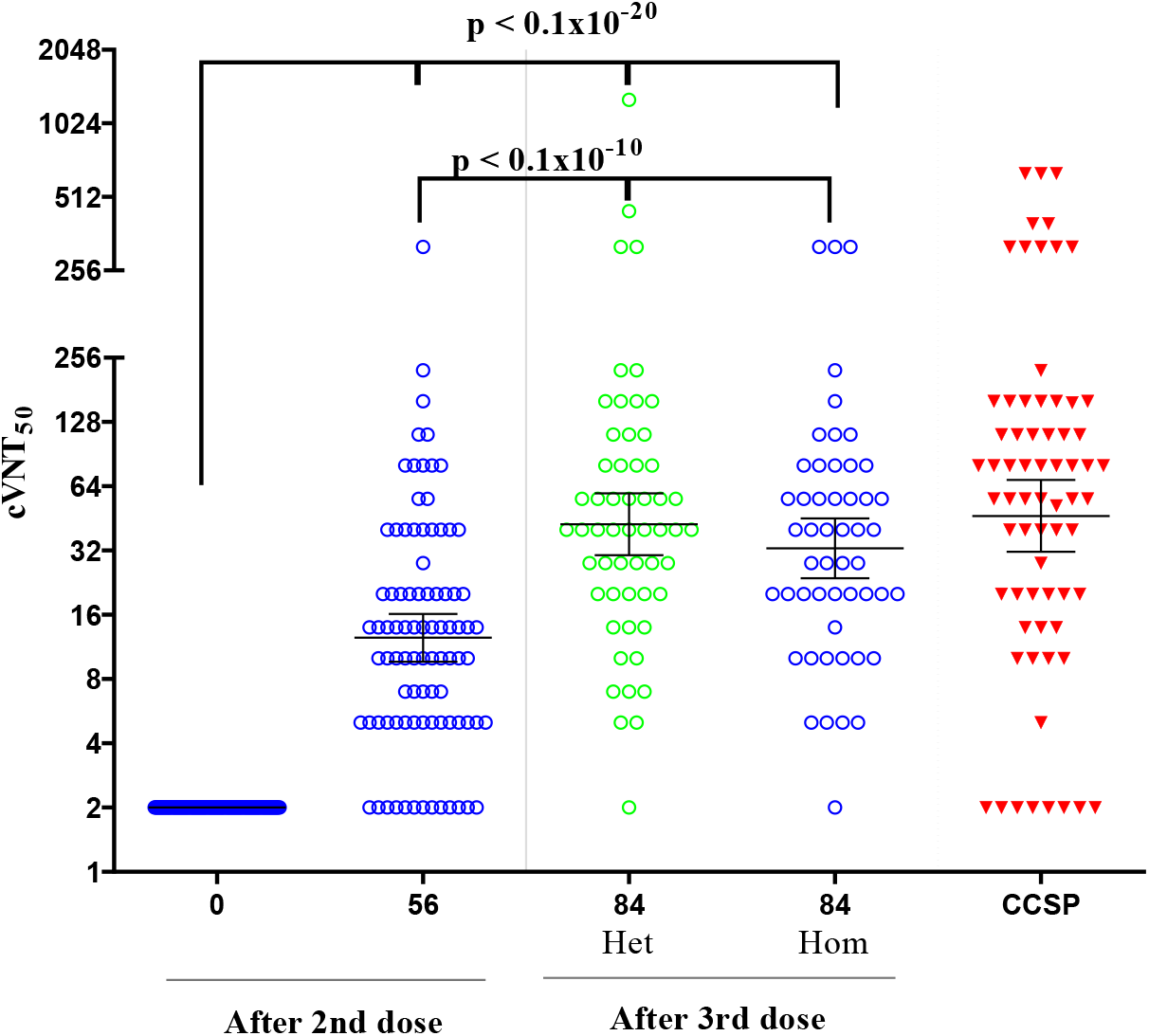
Neutralizing titre of SARS-CoV-2 (D614G) live-virus after two doses of SOBERANA 02 and a third homologous or heterologous dose (pooled analysis from phase I and phase IIa). FOOTNOTE: Subjects 19-80 years old received two doses (T0, T28) of SOBERANA 02 and a third dose (T56) homologous, (Hom: blue points) or heterologous (SOBERANA Plus, Het: green points). cVNT50: Conventional live-virus neutralization titre (GMT, IC 95%). CCSP: Cuban Convalescent Serum Panel (red triangles). p values represent the statistic differences with T0 or T56, as indicated, using paired Student t test with log-transformed variables.

The molecular neutralizing effect of anti-RBD IgG against phages displaying delta (δ)-RBD (L452R+T478K) compared to D614G**-**RBD was analysed in 16 serum samples from individuals vaccinated with the heterologous scheme. Figure 7 shows an mVNT_50_ GMT of 962.9 (95% IC 670.1; 1384) against phages displaying D614G-RBD and 384.1 (95% IC 262; 562.9) against δ-RBD phages, meaning a reduction of 2.5 fold the molecular neutralization capacity of the antibodies.

**Figure 7.**
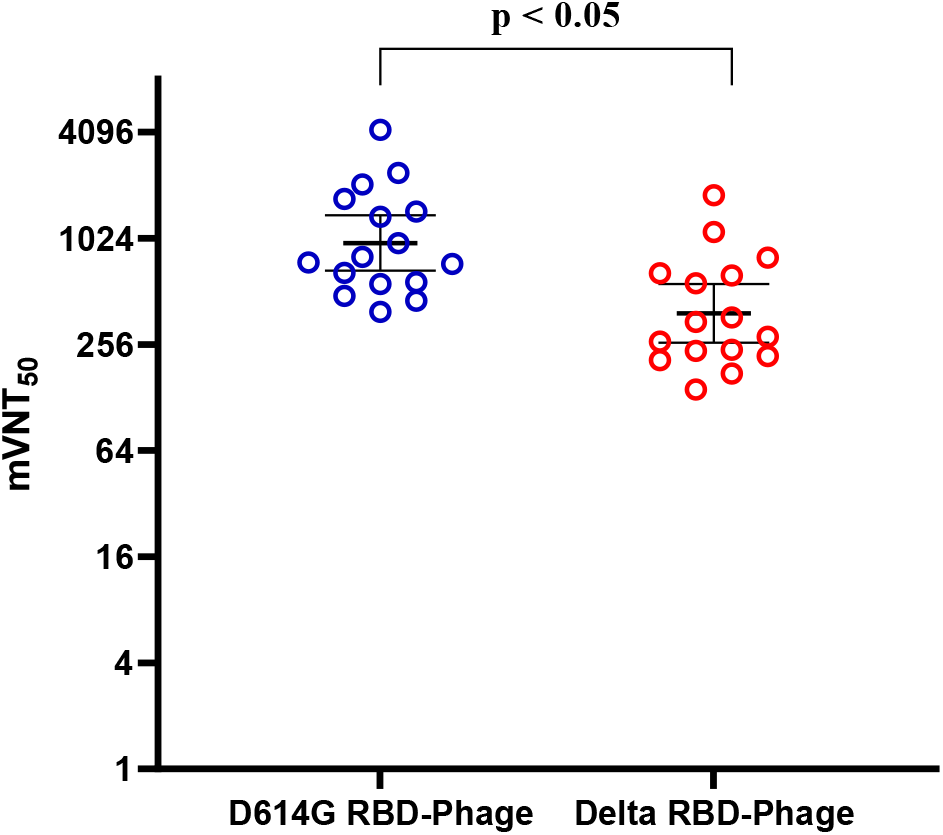
Anti-RBD IgG antibodies inhibit the interaction between the human ACE2 receptor and phages displaying D614G -RBD or δ-RBD variant. FOONOTE: Sera from 16 individuals vaccinated with heterologous schedule were tested (GMT, IC 95%). During the trial clinical trials, the predominant circulating strain was D614G. p value represents the statistic differences as indicated, using paired Student t test with log-transformed variables.

Both age subgroups (19-59 and 60-80) in phase IIa showed similar immunological responses (p≥0.05) after the third dose, the neutralizing antibodies titres were similar. Significant differences only were observed for the molecular neutralization titre (mVNT_50_) with higher values in the 19-59 years-group respect to 60-80 years-groups. (Supplemental Material, Appendix B, Table IX).

RBD-specific T cell response was assessed by IFN-γ and IL-4 expression in peripheral blood mononuclear cell (PBMC) in a subset of participants, as an indicative of Th1 or Th2 profile. After two doses of SOBERANA 02 (T56), the number of IFN-γ forming cells were statistically different (p<0,05) to baseline levels (T0) (Figure 8A). The number of IL-4 secreting cells did not increase (p>0,99) (Figure 8B) showing a classical profile of Th1 cellular immune response after two doses of SOBERANA 02. A significant increase of both IFN-γ producing cells (p<0,005) and IL-4 producing cells although significant (p<0,001) occurred after the third dose (T84), suggesting a mixture of Th1/Th2 profile. There were no differences between both alternative third doses (p>0,99).

**Figure 8.**
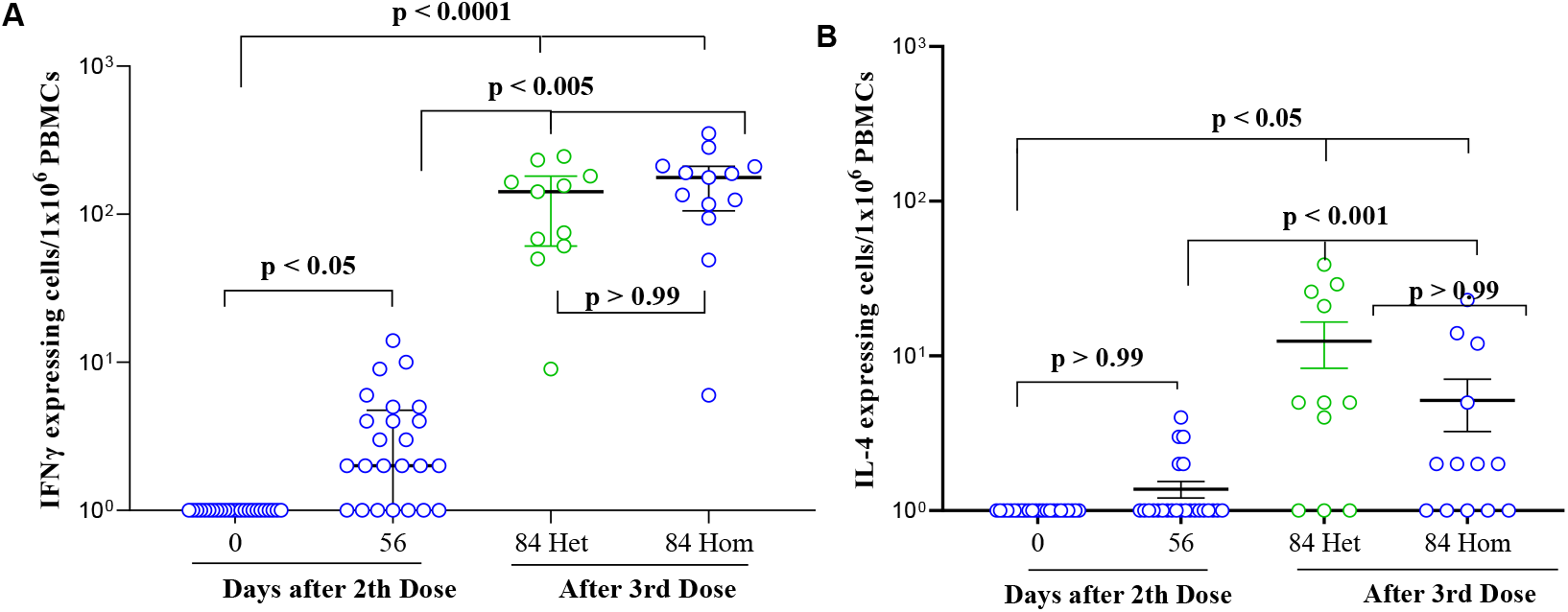
IFN-γ- and IL-4-secreting cells in peripheral blood mononuclear cells stimulated with RBD. FOONOTE: Subjects 19-80 years old received two doses (on days 0, 28; N=24) of SOBERANA 02 and a third dose (on day 56) homologous (SOBERANA 02, Hom: blue points, N=13) or heterologous (SOBERANA Plus, Het: green points; N=11). p value represents the statistic differences as indicated.

There was a good correlation among all variables (coefficients greater than 0·7, Supplemental Material, Appendix B, Table X). The likelihood ratio (using Bayes Factor) was used as Benefit-Risk index. In all considered scenarios, there is strong evidence for benefit, with a higher index for the heterologous scheme (Supplemental Material, Appendix B, Figure I).

## Discussion

Conjugate vaccines have been used for more than 30 years, mainly in children, for preventing bacterial infection diseases. Their induction of potent B and T immune responses, both endowed with immunological memory, marked a breakthrough in vaccinology [30]. SOBERANA 02 is an innovative conjugate vaccine in which the viral antigen RBD is conjugated to tetanus toxoid. As expected, SOBERANA 02 showed an excellent safety profile, in three doses homologous and heterologous schemes with SOBERANA Plus, with predominance of local over systemic AEs. The frequency of adverse events (50% in phase 1 and 31% in phase IIa), particularly the systemic AEs, is lower compared to anti-SARS-CoV-2 mRNA or adenovirus-vectored vaccines [31, 32, 33, 34, 35]. These results provide the first evidences of safety of SOBERANA 02-25 μg in three doses or in heterologous combination with SOBERANA Plus. In phase I, 25 µg-dose SOBERANA 02 was more immunogenic than 15 μg-dose; in consequence, after phase I interim analysis the high dose progressed to phase II trial.

Vaccine candidates eliciting similar or higher immune response as compared with convalescents serum panels have moved forward in clinical evaluation [35, 36, 37]. The pooled immune response data from phase I and phase IIa were compared with those from the Cuban Convalescent Serum Panel (CCSP). Two doses of SOBERANA 02-25 µg elicited similar immune response compared to the CCSP in terms of anti-RBD IgG titre and molecular inhibition of RBD:hACE2 interaction; however, elicited RBD antibodies showed a lower viral neutralization capacity. For this reason, the study incorporated a third dose. Both the homologous and the heterologous—incorporating SOBERANA Plus— three-dose schemes, increased the humoral immune response and titre of neutralizing antibodies. In both cases, neutralizing capacities were similar to the observed in convalescents.

The humoral immune response by age group was explored in phase IIa, including a small number of subjects aged 60-80; the results here-presented are encouraging in this age group that is severely affected by COVID-19 [38].

By mid-2021, the SARS-CoV-2 VOC δ became predominant worldwide, being 60% more transmissible than variant α [39] and reducing vaccine efficacy towards the onset of symptomatic disease [40, 41]. The predominant variant circulating in Havana at the moment of these studies was D614G, but it was replaced firstly by beta (March-June 2021) and later completely by delta (July-October 2021) [42]. We evaluated molecular neutralizing capacity (mVNT_50_) against VOC δ and found a decrease of 2.5-fold compared to molecular neutralizing capacity against D614G variant. This result is in correspondence with reports of three-to five-fold reduction in neutralization titres against VOC δ in respect to VOC α in sera form individuals immunized with mRNA vaccines or adenoviral-vectors [43]. Protection against the circulating VOCs will be addressed in the next clinical trials.

Specific T cell response plays an important role for anti-SARS-CoV-2 immunity [44; 45]. Our results demonstrate the activation of Th1 cellular immune pattern after two dose of SOBERANA 02, characterized by predominant IFN-γ over IL-4-secreting cells. After both types of third doses, both cytokines-secreting cells increased significantly, predominating IFN-γ secretion, and suggesting a balanced Th1/Th2 profile, contributing to the high increase in anti-RBD IgG levels.

The first heterologous scheme in COVID vaccine was reported for Sputnik V (two shots scheme with different adenoviral vector) [7]. Recent studies are evaluating heterologous booster effects of mRNA BNT162b2 in individuals previously vaccinated with adenoviral vaccines and other vaccine combinations as prime/boost heterologous strategy [46]. Our approach of heterologous 2+1 scheme was different. We focused in priming with two doses (0, 28 days) with RBD-tetanus toxoid conjugate vaccine SOBERANA 02 for inducing specific humoral and cellular immune response favoured by the multiepitopic presentation of RBD [28, 29]; followed by a third dose (on day 56) with SOBERANA Plus (dimeric-RBD/alumina), changing the RBD epitope presentation to the immune system. Although the sample size did not allow for statistical comparisons between heterologous and homologous schemes, these results encouraged us to move to phase IIb and phase III trial with the heterologous scheme.

Both three-dose schemes were equally safe; in contrast to a recent report where heterologous boost of mRNA vaccine in individuals previously vaccinated with ChAdOx1 (ChAd) increased systemic reactogenicity as compared with homologous boost with ChAdOx1 (ChAd) [47].

The main limitation of our study is its open label design; the lack of a control-placebo group precludes the comparison with unvaccinated subjects.

In conclusion, SOBERANA 02 is safe, well tolerated and immunogenic in adult aged 19-80 years. Application of a heterologous scheme with SOBERANA Plus increased the immune response with excellent safety profile, reinforcing the induction of memory cells. These results pave the way for further evaluation of the heterologous scheme in phase IIb and phase III clinical trials.

## Data Availability

All data produced in the present study are available upon reasonable request to the authors

https://www.finlay.edu.cu/blog/category/sala-cientifica/

## Acknowledgments

We especially thank all the volunteers who participated in these first clinical trials. Also, to Dr. Lila Castellanos for scientific advice and language corrections.

