## Supplemental Material for "Safety and immunogenicity of anti-SARS CoV-2 vaccine SOBERANA 02 in homologous or heterologous scheme"

**INDEX**

**Appendix A**

A-1. Selection criteria for phase I clinical trial

A-2. Selection criteria for phase IIa clinical trial

A3- Outcomes.

**Appendix B.**

Table I. Frequency of subjects with adverse events, during phase I and phase IIa clinical trials.

Table II. Characterization of adverse events

Table III: Frequency of subjects with adverse events by dose, during phase I and phase IIa clinical trials.

Table IV. Haematology and Blood Chemistry

Table V: Immune response elicited after two doses of SOBERANA02 and third homologous or heterologous dose, during phase I and phase IIa.

Table VI. Kinetic of immune response after two doses of SOBERANA 02 and a third homologous or heterologous dose (pooled analysis from phase I and phase IIa)

Table VII. Anti-RBD IgG antibodies inhibit the interaction between RBD and human ACE2 receptor after two doses of SOBERANA 02 and a third homologous or heterologous dose (pooled analysis from phase I and phase IIa).

Table VIII. Neutralizing titre of SARS-CoV-2 live-virus after two doses of SOBERANA 02 and a third homologous or heterologous dose (pooled analysis from phase I and phase IIa).

Table IX. Comparison of immune response induced by age groups after three doses (only Phase IIa)

Table X. Bivariate correlation after three doses between immune variables (pooled analysis from Phase I and Phase IIa)

Figure I. Risk-Benefit Balance.

**Appendix C. Immunological Techniques**

C.1 Anti-RBD IgG response

C.2 Molecular virus neutralization tests

C.3. Phages construction

C.4. Molecular virus neutralization test with phage-displayed RBDs

C.5 Conventional virus neutralization test

C.6 Specific T-cell response

**Appendix A.**

**A-1. Selection criteria for phase I clinical trial**

Inclusion criteria:

1. Subjects who give written informed consent to participate in the study.

2. Subjects aged 19– 59 years.

3. Women of childbearing potential, using safe contraceptive methods during the study.

4. Physical examination: normal or without clinically significant alterations.

5. Laboratory results within or outside the range of reference values but not clinically significant.

Exclusion criteria:

1. acute febrile or infectious disease in the 7 days prior to the administration of the vaccine or at the time of its application.

2. antimicrobial treatment in the 7 days prior to the administration of the vaccine.

3. Weight Loss (BMI <18.5) and obesity (BMI ≥ 30.0)

4. chronic non-transmissible diseases not controlled according to clinical or laboratory criteria (eg bronchial asthma, chronic obstructive pulmonary disease, diabetes mellitus, thyroid diseases, ischemic heart disease, arterial hypertension, psychiatric, neurological, hematologic disease).

5. congenital or acquired immune system disease.

6. history of unresolved neoplastic disease.

7. personal history of liver or kidney failure.

8. history of substance abuse within the past 30 days or substance addictive illness, except withdrawal and smoking.

9. diminished mental faculties for decision making.

10. history of severe allergic disease (anaphylactic shock, angioneurotic oedema, glottis oedema, severe urticaria).

11. history of hypersensitivity to thiomersal or to some of the components of the formulation.

12. history of SARS and COVID-19 who meet any of the following criteria: a) Previous or current history of SARS-CoV-2 infection. b) Be declared in the category of contact or suspect at the time of inclusion c) positive test for anti-SARS-CoV-2 Antibodies. d) Positive PCR at the time of inclusion.

13. Participation in another clinical trial in the last 3 months.

14. Application of vaccines containing tetanus toxoid in the last 3 months.

15. Application of other vaccines in the last 30 days.

16. Treatment with immunomodulators in the last 30 days

17. Transfusion of blood or blood products in the last 3 months.

18. Subjects with difficulties in attending the planned follow-up consultations.

19. Splenectomy or splenic dysfunction.

20. Pregnancy, puerperium or breastfeed

21. Subjects with tattoos in the deltoid region on both arms.

22. Subjects with positive results for HIV, hepatitis B surface antigen, hepatitis C antibody, and syphilis positive serology.

**A-2. Selection criteria for phase IIa clinical trial**

Inclusion criteria:

1. Subjects who give written informed consent to participate in the study.

2. Subjects aged 19–80 years.

3. Women of childbearing age using contraceptive methods during the study.

4. Physical examination: normal or without clinically significant alterations.

5. Laboratory results within or outside the range of reference values but not clinically significant (for stage IIa)

Exclusion criteria:

1. acute febrile or infectious disease in the 7 days prior to the administration of the vaccine or at the time of its application.

2. antimicrobial treatment in the 7 days prior to the administration of the vaccine.

3. Low weight (BMI <18.5) and obesity (BMI ≥ 34.9).

4. non-transmissible chronic diseases not controlled according to clinical or laboratory criteria (eg bronchial asthma, chronic obstructive pulmonary disease, diabetes mellitus, thyroid diseases, ischemic heart disease, arterial hypertension, psychiatric disease haemotologic disease).

5. congenital or acquired immune system disease.

6. history of unresolved neoplastic disease.

7. personal history of liver or kidney failure.

8. history of substance abuse during the last 30 days or addictive illness to toxic substances, except if the subject is in abstinence, in the case of alcoholics, and smoking.

9. diminished mental faculties for decision making.

10. history of severe allergic disease (anaphylactic shock, angioneurotic oedema, glottis oedema, severe urticaria).

11. history of hypersensitivity to thiomersal or to some of the components of the formulation.

12. history of SARS and COVID-19 who meet any of the following criteria: a) Previous or current history of SARS-CoV-2 infection. b) Be declared in the category of contact or suspect at the time of inclusion. c) Subject with positive test for Anti-SARS-CoV-2 Antibodies. d) Subject with positive PCR at the time of inclusion.

13. Participation in another clinical trial in the last 3 months.

14. Application of vaccines containing tetanus toxoid in the last 3 months.

15. Application of other vaccines in the last 30 days.

16. Treatment with immunomodulators in the last 30 days

17. Transfusion of blood or blood products in the last 3 months.

18. Subjects with difficulties in attending the planned follow-up consultations.

19. Splenectomy or splenic dysfunction.

20. Pregnancy, puerperium or breastfeed

21. Subjects with tattoos in the deltoid region on both arms.

22. Subjects with positive results for HIV, Hepatitis B Surface Antigen, Hepatitis C Antibody and syphilis positive serology.

23. Women with a positive pregnancy test.

**A3- Outcomes.**

*Primary outcome(s):*

1) Serious Adverse Events-SAE

Measured as: a) Occurrence of the SAE (Yes, No), b) Duration (Time since the beginning until the end of the event); c) Description of the event; d) Result (Recovered, Recovered with squeals, Persists, Death, Unknown); e) Causality association (Consistent causal relation to immunization, Inconsistent causal relation to immunization, Indeterminate, Unclassifiable) Measurementsdaily for 28 days after each dose.

*Secondary outcomes:*

1) Solicited Local and systemic Adverse Events (AE);

2) Unsolicited Adverse Events (AE);

- Measure as: Description of the AE (name of the event), Duration (Time from start date until end date of event), -Intensity of the AE (mild, moderate, severe), -Severe (Serious, not serious), -Result (Recovered, Recovered with sequelae, Persists, Death, Unknown), -Causality (causal association consistent with vaccination, Undetermined, causal association inconsistent with vaccination, not classifiable).

- Measurement time: daily for 7 days after each dose for the solicited AE.

- Measurement time: daily for 28 days after each dose for the unsolicited AE.

*Secondary outcomes for immunogenicity:*

4) Specific anti-RBD IgG antibodies and percentage of subjects with seroconversion 4-fold to pre-vaccination. Measurement on days 0, 14, 28, 42, 56, 70; 84.

5) Conventional neutralizing antibody titre: Measurement on days 0, 56 and 84

6) % ACE2-RBD inhibition: Measurement on days 0, 14, 42, 56, 70, 84.

*Others:*

7) Molecular neutralizing titre using WT RBD (measurement time: Day 14, 42, 56, 70, 84) and using mutant variant Delta-L452R+T478K. (Measurement on days 84, only in phase IIa)

8) RBD-specific T-cells responses producing IFN- γ and TNF- α, (Measurement on days 0, 56, 84 only in phase IIa)

**Appendix B.**

**Table I. Frequency of subjects with adverse events, during phase I and phase IIa clinical trials.**

|  | | | | Phase I | | Phase IIa |
| --- | --- | --- | --- | --- | --- | --- |
|  |  |  |  | Arm 1:  SOBERANA 02-15 µg N=20 | Arm 2:  SOBERANA 02-25 µg N=20 | SOBERANA02-25 µg N=100 |
|  |  |  |  | **Freq. (%)** | **Freq. (%)** | **Freq. (%)** |
| N | | | | 20 (100) | 20 (100) | 100 (100) |
| Subjects With AE | | | | 6 (30.0) | 10 (50.0) | 32 (32) |
| Solicited | Local | | Injection-site pain | 0 | 3 (15.0) | 22 (22) |
|  |  |  | Erythema | 0 | 0 | 4 (4.0) |
|  |  |  | Local warm | 0 | 0 | 4 (4.0) |
|  |  |  | Induration | 0 | 0 | 3 (3.0) |
|  |  |  | Swelling | 0 | 0 | 3 (3.0) |
|  | Systemic | | General discomfort | 0 | 0 | 5 (5.0) |
|  |  |  | Rash | 0 | 0 | 1 (1.0) |
| Unsolicited | Systemic | | Acute bronchitis | 1 (5.0) | 0 | 0 |
|  |  |  | Cellulitis | 1 (5.0) | 0 | 0 |
|  |  |  | Rhinitis | 1 (5.0) | 1 (5.0) | 0 |
|  |  |  | Cough | 1 (5.0) | 0 | 1 (1.0) |
|  |  |  | Headache | 0 | 1 (5.0) | 5 (5.0) |
|  |  |  | Dengue | 0 | 1 (5.0) | 0 |
|  |  |  | High blood pressure | 3 (15.0) | 5 (25.0) | 4 (4.0) |
|  |  |  | Food poisoning | 1 (5.0) | 0 | 0 |
|  |  |  | Tachycardia | 0 (0.0) | 1 (5.0) | 0 |
|  |  |  | Upper Acute respiratory infection | 2 (10.0) | 0 | 5 (5.0) |
|  |  |  | Herpes zoster | 0 | 0 | 1 (1.0) |
|  |  |  | Itchy throat | 0 | 0 | 1 (1.0) |
|  |  |  | Urinary sepsis | 0 | 0 | 1 (1.0) |
|  |  |  | Asthenia | 0 | 0 | 2 (2.0) |
|  |  |  | Drowsiness | 0 | 0 | 1 (1.0) |
|  |  |  | Bilateral orchiepididymitis | 0 | 0 | 1 (1.0) |
|  |  |  | Diarrhea | 0 | 0 | 1 (1.0) |
| Total of AEs | | | | 10 | 13 | 72 |
| Number of AE by subject | | Mean ± SD | | 0.5 ± 0.9 | 0.6 ± 0.8 | 0.7 ± 1.5 |
|  |  | Median ± IQR | | 0 ± 1 | 0.5 ± 1 | 0 ± 1 |
|  |  | (Min; Max.) | | (0; 3) | (0; 3) | (0; 7) |

**Table II. Characterization of adverse events**

| **Global characterization** | | **Phase I** | | **Phase IIa** |
| --- | --- | --- | --- | --- |
|  |  | Arm 1:  SOBERANA 02-15 µg N=20 | Arm 2:  SOBERANA 02-25 µg N=20 | SOBERANA 02-25 µg  N=100 |
|  |  | **Freq.**  **%** | **Freq.**  **%** | **Freq.**  **%** |
| Total of adverse events | | 10 (100) | 13 (100) | 72 (100) |
| Intensity | Mild (Grade 1) | 7 (70.0) | 11 (84.6) | 65 (90.3) |
|  | Moderate (Grade 2) | 2 (20.0) | 1 (7.7) | 4 (5.6) |
|  | Severe (Grade 3) | 1* (10.0) | 1** (7.7) | 3***(4.2) |
| Severity | Serious | 0 | 1* (7.7) | 2* (2.8) |
|  | No Serious | 10 (100) | 12 (92.3) | 70 (97.2) |
| Causal relationship | Consistent with vaccination  (Vaccine related) | 0 | 3 (23.1) | 58 (80.6) |
|  | Consistent with vaccination (Conditions inherent to the vaccinated) | 2 (20.0) | 7 (53.8) | 7 (9.7) |
|  | Inconsistent with vaccination | 8 (80.0) | 3 (23.1) | 7 (9.7) |
| Result | Recovered | 10 (100) | 12 (92.3) | 71 (98.6) |
|  | Recovered with sequels | 0 | 1^&^ (7.7) | 1^#^ (1.4) |
| Type | Local | 0 | 4 (30.8) | 43 (59.7) |
|  | Systemic | 10 (100) | 9 (69.2) | 29 (40.3) |
| Solicited | Solicited | 0 | 4 (30.8) | 49 (68.1) |
|  | Unsolicited | 10 (100) | 9 (69.2) | 23 (31.9) |
| Duration (hours) | ≤ 24 hours | 3 (30.0) | 9 (69.2) | 56 (77.8) |
|  | 24-48 hours | 3 (30.0) | 1 (7.7) | 7 (9.7) |
|  | 48-72 hours | 1 (10.1) | 1 (7.7) | 4 (5.6) |
|  | > 72 hours | 3 (30.0) | 0 | 5 (6.9) |
|  | Unspecified | 0 | 2 (15.4) | 0 |
| *Grade 3: *Food poisoning; *Dengue*^&^*; * Erythema, induration, Bilateral orchiepididymitis*  *^#^: Herpes zoster* | | | | |

**Table III: Frequency of subjects with adverse events by dose, during phase I and phase IIa clinical trials.**

|  | **Phase I.** | | **Phase IIa** |
| --- | --- | --- | --- |
|  | Arm 1:  SOBERANA 02-15 µg  N=20 | Arm 2:  SOBERANA 02-25 µg N=20 | SOBERANA 02-25 µg;  N=100 |
| **Overall AEs** | | | |
| Subjects with AE | 6 (30%) | 10 (50%) | 32 (32%) |
| 1^st^ dose | 6/20 (30.0%) | 7/20 (35.0%) | 27/100 (27.0%) |
| 2^nd^ dose | 2/20 (10.0%) | 3/19 (15.8%) | 9/97 (9.3%) |
| 3^rd^ dose-Hom | 0 | 0 | 4/47 (8.5%) |
| 3^rd^ dose-Het | 0 | 0 | 2/49 (4.1%) |
| **Subjects with solicited AEs** | | | |
| Any | 0 | 3 (15.0%) | 25 (25.0%) |
| 1^st^ dose | 0 | 1/20 (5.0%) | 21/100 (21.0%) |
| 2^nd^ dose | 0 | 2/19 (10.5%) | 8/97 (8.2%%) |
| 3^rd^ dose-Hom | 0 | 0 | 4/47 (8.5%) |
| 3^rd^ dose-Het | 0 | 0 | 2/49 (4.1%) |
| **Subjects with solicited systemic AEs** | | | |
| Any | 0 | 0 | 6 (6%) |
| 1^st^ dose | 0 | 0 | 3/100 (3.0%) |
| 2^nd^ dose | 0 | 0 | 2/97 (2.1%) |
| 3^rd^ dose-Hom | 0 | 0 | 0 |
| 3^rd^ dose-Het | 0 | 0 | 1/49 (2.0%) |
| General discomfort | 0 | 0 | 5 (5.0%) |
| 1^st^ dose | 0 | 0 | 2/100 (2.0%) |
| 2^nd^ dose | 0 | 0 | 2/97 (2.1%) |
| 3^rd^ dose-Hom | 0 | 0 | 0 |
| 3^rd^ dose-Het | 0 | 0 | 1/49 (2.0%) |
| Rash | 0 | 0 | 1 (1.0%) |
| 1^st^ dose | 0 | 0 | 1/100 (1.0) |
| 2^nd^ dose | 0 | 0 | 0 |
| 3^rd^ dose-Hom | 0 | 0 | 0 |
| 3^rd^ dose-Het | 0 | 0 | 0 |
| **Subjects with solicited AEs** | | | |
| Any | 0 | 3 (15%) | 22 (22.0%) |
| 1^st^ dose | 0 | 1/20 (5.0%) | 18/100 (18.0%) |
| 2^nd^ dose | 0 | 2/19 (10.5%) | 7/97 (7.2%) |
| 3^rd^ dose-Hom | 0 | 0 | 4/47 (8.5%) |
| 3^rd^ dose-Het | 0 | 0 | 2/49 (4.1%) |
| Injection-site pain | 0 | 3 (15%) | 22 (22%) |
| 1^st^ dose | 0 | 1/20 (5.0%) | 17/100 (17.0%) |
| 2^nd^ dose | 0 | 2/19 (10.5%) | 6/97 (6.2%) |
| 3^rd^ dose-Hom | 0 | 0 | 4/47 (8.5%) |
| 3^rd^ dose-Het | 0 | 0 | 2/49 (4.1%) |
| Erythema | 0 | 0 | 4 (4.0%) |
| 1^st^ dose | 0 | 0 | 2/100 (2.0%) |
| 2^nd^ dose |  |  | 0 |
| 3^rd^ dose-Hom | 0 | 0 | 2/47 (4.2%) |
| 3^rd^ dose-Het | 0 | 0 | 0 |
| Local Warm | 0 | 0 | 4 (4.0%) |
| 1^st^ dose | 0 | 0 | 0 |
| 2^nd^ dose | 0 | 0 | 1/97 (1.0%) |
| 3^rd^ dose-Hom | 0 | 0 | 3/47 (6.4%) |
| 3^rd^ dose-Het | 0 | 0 | 0 |
| Induration | 0 | 0 | 3 (3.0%) |
| 1^st^ dose | 0 | 0 | 0 |
| 2^nd^ dose | 0 | 0 | 1/97 (1.0%) |
| 3^rd^ dose-Hom | 0 | 0 | 2/47 (4.2%) |
| 3^rd^ dose-Het | 0 | 0 | 0 |
| Swelling | 0 | 0 | 3 (3.0%) |
| 1^st^ dose | 0 | 0 | 0 |
| 2^nd^ dose | 0 | 0 | 1/97 (1.0%) |
| 3^rd^ dose-Hom | 0 | 0 | 2/47 (4.2%) |
| 3^rd^ dose-Het | 0 | 0 | 0 |
| **Subjects with unsolicited AEs** | | | |
| Any | 6 (30.0%)* | 7 (35.0%)** | 17 (17.0%)*** |
| 1^st^ dose | 6/20 (30.0%) | 6/20 (30.0%) | 13/100 (13.0%) |
| 2^nd^ dose | 2/20 (10.0) | 1/19 (5.3%) | 2/97 (2.1%) |
| 3^rd^ dose-Hom | 0 | 0 | 1/47 (2.1%) |
| 3^rd^ dose-Het | 0 | 0 | 2/49 (4.1%) |
| *Hom: Homologous third dose; Het: Heterologous third dose*  ** Acute Bronchitis (1/20: 2nd dose); cellulitis (1/20: 1st dose), High blood pressure (3/20: 1st dose); Food poisoning (1/20: 1st dose); rhinitis (1/20: 2nd dose); nasal secretion (1/20: 1st dose); acute sinusitis (1/20: 1st dose); dry cough (1/20: 1st dose)*  *** Headache (1/20: 1st dose), High blood pressure (5/20: 1st dose); Dengue (1/20: 1st dose); rhinitis (1/19: 2nd dose), tachycardia (1/20: 1st dose)*  **** Asthenia (1/97: 2nd dose; 1/49; 3rd dose-Het); Headache (3/100: 1st dose; 1/47: 3rd dose-Hom; 1/49: 3rd dose-Het); diarrhea (1/49: 3rd dose-Het); pharyngeal burning (1/100: 1st dose); herpes zoster (1/97: 2nd dose); High blood pressure (4/100: 1st dose); Acute respiratory infection (5/100: 1st dose); bilateral orchiepididymitis (1/100: 1st dose); urinary sepsis (1/100: 1st dose); somnolence (1/97: 2nd dose)* | | | |

**Table IV. Haematology and Blood Chemistry**

|  | | | Phase I | | Phase IIa | |
| --- | --- | --- | --- | --- | --- | --- |
|  | | | Arm 1:  SOBERANA 02-  15 µg | Arm 2: SOBERANA 02-25 µg | SOBERANA 02-25 µg | |
|  |  |  |  |  | Third Dose-Hom* | Third-Dose  Het* |
| N | | | 20 | 20 | 44 | 47 |
| **Haemoglobin** | Pre | Median (25-75th) | 154 (138; 160) | 146 (133; 156) | 136 (128; 148) | 139 (131; 157) |
|  |  | (Range) | (129;169) | (120; 164) | (113; 158) | (110; 180) |
|  | Post | Median (25-75th) | 154 (138; 159) | 148 (134; 158) | 140 (130; 150) | 147 (133; 157) |
|  |  | (Range) | (129; 167) | (119; 135) | (106; 166) | (117; 177) |
| **Haematocrit** | Pre | Median  (25-75th) | 41.8  (38.2; 43.4) | 40.0  (36.9; 42.7) | 39.1  (36.4; 41.4) | 39.1  (37.1; 42.9) |
|  |  | (Range) | (36.0; 46.8) | (33.7; 45.4) | (31.9; 44..9) | (33.0; 48.5) |
|  | Post | Median (25-75th) | 41.4  (37.7; 41.4) | 39.7  (36.2; 42.2) | 40.0  (37.9; 43.2) | 41.8  (38.5; 44.7) |
|  |  | (Range) | (36.0; 46.2) | (32.8; 44.9) | (31.9; 49.1) | (33.9; 49.0) |
| **Platelets** | Pre | Median (25-75th) | 223 (189; 257) | 251 (203; 297) | 239 (210; 273) | 234 (205; 265) |
|  |  | (Range) | (137; 339) | (132; 382) | (162; 341) | (167; 423) |
|  | Post | Median (25-75th) | 215 (189; 261) | 245 (214; 297) | 234 (199; 268) | 248 (211; 278) |
|  |  | (Range) | (156; 320) | (132; 363) | (161; 373) | (170; 354) |
| **Leukocytes** | Pre | Median (25-75th) | 7.2 (6.4; 8.2) | 7.4 (5.6; 8.0) | 6.4 (5.4; 7.5) | 6.8 (6.1; 7.7) |
|  |  | (Range) | (5.3; 13.5) | (4.2; 10.8) | (4.5; 10.7) | (5.0; 9.4) |
|  | Post | Median (25-75th) | 7.2 (6.3; 8.5) | 7.0 (5.6; 8.0) | 5.8 (4.9; 6.7) | 5.5 (5.1; 6.9) |
|  |  | (Range) | (5.3; 11.8) | (4.2; 10.8) | (3.2; 8.6) | (4.1; 8.7) |
| **Lymphocytes** | Pre | Median (25-75th) | 2.5 (2.2; 3.1) | 2.2 (2.0; 2.6) | 2.2 (1.8; 2.7) | 2.2 (1.8; 2.5) |
|  |  | (Range) | (1.4; 3.9) | (1.4; 3.5) | (0.1; 3.6) | (1.5; 3.9) |
|  | Post | Median (25-75th) | 2.4 (1.8; 3.0) | 1.9 (1.7; 2.3) | 2.1 (1.7; 2.6) | 2.0 (1.7; 2.2) |
|  |  | (Range) | (1.5; 3.8) | (1.4; 2.3) | (1.3; 3.4) | (1.3; 4.2) |
| **Monocytes** | Pre | Median (25-75th) | 0.50  (0.45; 0.62) | 0.49  (0.40; 0.61) | 0.40  (0.35; 0.46) | 0.47  (0.40; 0.56) |
|  |  | (Range) | (0.27; 0.74) | (0.26; 0.73) | (0.24; 0.78) | (0.28; 1.00) |
|  | Post | Median (25-75th) | 0.53  (0.47; 0.64) | 0.46  (0.35; 0.62) | 0.36  (0.31; 0.44) | 0.42  (0.35; 0.51) |
|  |  | (Range) | (0.30; 1.04) | (0.24; 0.97) | (0.19; 0.77) | (0.19; 0.80) |
| **Eosinophils** | Pre | Median (25-75th) | 0.15  (0.10; 0.23) | 0.14  (0.10; 0.19) | 0.10  (0.07; 0.22) | 0.14  (0.10; 0.23) |
|  |  | (Range) | (0.04; 0.71) | (0.03; 0.39) | (0.00; 0.54) | (0.03; 0.73) |
|  | Post | Median (25-75th) | 0.20  (0.11; 0.34) | 0.13  (0.08; 0.23) | 0.16  (0.11; 0.24) | 0.16  (0.11; 0.24) |
|  |  | (Range) | (0.05; 0.95) | (0.02; 0.49) | (0.04; 1.01) | (0.06; 0.68) |
| **Basophils** | Pre | Median  (25-75th) | 0.02  (0.02; 0.04) | 0.02  (0.02; 0.03) | 0.02  (0.02; 0.03) | 0.02 (0.02; 0.04) |
|  |  | (Range) | (0.01; 0.07) | (0.01; 0.04) | (0.00; 0.07) | (0.01; 0.08) |
|  | Post | Median  (25-75th) | 0.02  (0.02; 0.04) | 0.03  (0.02; 0.03) | 0.02  (0.02; 0.03) | 0.02  (0.02; 0.04) |
|  |  | (Range) | (0.01; 0.07) | (0.01; 0.04) | (0.00; 0.07) | (0.01; 0.08) |
| **Neutrophils** | Pre | Median (25-75th) | 3.6 (3.4; 4.6) | 4.0 (3.5; 5.0) | 3.3 (2.8; 4.4) | 3.8 (3.2; 4.4) |
|  |  | (Range) | (3.0; 8.2) | (2.5; 6.5) | (1.6; 7.3) | (1.6; 7.3) |
|  | Post | Median (25-75th) | 4.2 (3.1; 4.9) | 4.2 (3.1; 5.0) | 3.0 (2.4; 3.4) | 3.1 (2.6; 3.8) |
|  |  | (Range) | (2.2; 7.7) | (2.3; 7.5) | (0.9; 5.8) | (1.5; 4.8) |
| **Glucose** | Pre | Median (25-75th) | 4.6 (4.4; 5.0) | 4.6 (4.3; 4.9) | 4.0 (3.5; 4.4) | 3.8 (3.5; 4.2) |
|  |  | (Range) | (3.9; 5.4) | (3.8; 5.1) | (2.8; 11.7) | (2.7; 6.0) |
|  | Post | Median (25-75th) | 3.7 (3.2; 4.2) | 4.0 (3.8; 4.5) | 4.2 (4.0; 4.6) | 4.2 (4.0; 4.6) |
|  |  | (Range) | (2.3; 4.9) | (3.0; 4.8) | (3.0; 14.0) | (3.2; 5.9) |
| **Creatinine** | Pre | Median (25-75th) | 86 (72; 95) | 77 (68; 93) | 73 (62; 90) | 80 (67; 90) |
|  |  | (Range) | (58; 114) | (50; 109) | (55; 115) | (53; 113) |
|  | Post | Median (25-75th) | 86 (69; 95) | 78 (68; 95) | 74 (62; 92) | 80(69; 93) |
|  |  | (Range) | (59; 114) | (56; 109) | (53; 121) | (51; 103) |
| **Uric acid** | Pre | Median (25-75th) | 339 (270; 381) | 272 (194; 333) | 260 (219; 322) | 294 (234; 340) |
|  |  | (Range) | (175; 480) | (164; 423) | (136; 403) | (169; 488) |
|  | Post | Median (25-75th) | 316 (265; 370) | 272 (185; 336) | 254 (222; 307) | 273 (232; 342) |
|  |  | (Range) | (175; 480) | (164; 388) | (107; 436) | (166; 572) |
| **ASAT** | Pre | Median (25-75th) | 20 (17; 26) | 19 (15; 23) | 18 (14; 23) | 19 (18; 22) |
|  |  | (Range) | (9; 36) | (8; 44) | (9; 47) | (13; 28) |
|  | Post | Median (25-75th) | 19 (17; 27) | 18 (15; 21) | 16 (13; 21) | 18 (14; 20) |
|  |  | (Range) | (9; 34) | (11; 28) | (8; 41) | (9; 24) |
| **ALAT** | Pre | Median (25-75th) | 26 (19; 46) | 19 (15; 25) | 14 (10; 20) | 16 (12; 25) |
|  |  | (Range) | (6; 91) | (8; 50) | (7; 48) | (9; 64) |
|  | Post | Median (25-75th) | 22 (15; 29) | 22 (11; 26) | 16 (13; 21) | 20 (16; 24) |
|  |  | (Range) | (6; 82) | (8; 50) | (8; 41) | (11; 40) |

**Hom: Homologous third dose; Het: Heterologous third dose*

**Table V: Immune response elicited after two doses of SOBERANA02 and third homologous or heterologous dose, during phase I and phase IIa.**

|  |  | **Phase I** | | | | | | | |  |
| --- | --- | --- | --- | --- | --- | --- | --- | --- | --- | --- |
|  |  | **Arm 1: SOBERANA 02-15** **µg** | | | | **Arm 2: SOBERANA 02-25 µg** | | | | **CCSP** |
|  |  | T0 | Post-2^nd^ dose | Post-3^rd^ dose-Hom | Post-3^rd^ dose-Het | T0 | Post-2^nd^ dose | Post-3^rd^ dose-Hom | Post-3^rd^ dose-Het | - |
| **Anti-RBD IgG AU/mL** | Seroconv. n/N; (%) | -- | 15/20 (75.0) | 6/7 (85.7) | 7/7 (100%) | -- | 15/19 (78.9) | 6/7 (85.7) | 9/9 (100.0) | - |
|  | 95% CI | -- | 50.9; 91.3 | 42.1; 99.6 | 91.6; 100 | -- | 54.4; 93.6 | 42.1; 99.6 | 93.5; 100 | - |
|  | Median | 3.1 | 25.9 | 61.2 | 59.9* | 3.2 | 40.3 | 208.2* | 165.2* | 50.8 |
|  | 25^th^-75^th^ | 3.1; 3.1 | 14.9; 39.5 | 14.9; 74.3 | 38.3; 169.2 | 3.1; 3.4 | 18.5; 102.9 | 33.8; 291.1 | 58.9; 633.3 | 23.8; 94.0 |
| **% Inh RBD:hACE2** | Median | 2.9 | 11.2 | 18.9 | 37.7* | 7.8 | 27.4 | 60.9* | 89.2* | 32.0 |
|  | 25^th^-75^th^ | 1.7; 6.8 | 6.4; 24.7 | 0.0; 51.3 | 22.1; 86.3 | 4.2; 9.4 | 10.2; 58.0 | 11.9; 87.6 | 57.2; 94.2 | 16.6; 62.3 |
| **mVNT_50_** | GMT | -- | 15.6 | 22.6 | 104.0*** | -- | 45.2 | 94.5* | 340.0*** | 41.8 |
|  | 95% CI | -- | 9.1; 26.9 | 4.1; 124.7 | 31.6; 342.7 | -- | 21.1; 96.9 | 18.5; 481.2 | 125.8; 918.5 | 27.7; 63.2 |
| **cVNT_50_** | GMT | 0 | 5.8 | 15.4 | 17.8 | 0 | 21.7 | 24.2* | 65.6* | 46.4 |
|  | 95% CI | 0 | 4.5; 7.4 | 9.0; 26.1 | 6.2; 50.7 | 0 | 7.8; 60.3 | 9.0; 65.3 | 22.0; 195.8 | 31.5; 68.4 |
|  |  | **Phase IIa SOBERANA 02-25 µg CCSP** | | | | | | |  |  |
|  |  | T0 | Post-2^nd^ dose | Post-3^rd^ dose-Hom | Post-3^rd^ dose-  Het |  | - |  |  |  |
| **Anti-RBD IgG AU/mL** | Seroconv. | -- | 71/94 (75.5) | 44/44 (100.0) | 48/49 (98.0) |  | - |  |  |  |
|  | 95% CI | -- | 65.6; 83.8 | 98.7; 100.0 | 89.1; 99.9 |  | - |  |  |  |
|  | Median | 1.9 | 23.2 | 80.9*** | 98.7*** |  | 50.8 |  |  |  |
|  | 25^th^-75^th^ | 1.9; 1.9 | 8.5; 69.9 | 39.9; 194.1 | 66.7; 256.7 |  | 23.8; 94.0 |  |  |  |
| **% Inh RBD:hACE2** | Median | 4.0 | 29.2 | 71.8*** | 79.7*** |  | 32.0 |  |  |  |
|  | 25^th^-75^th^ | 1.3; 6.6 | 13.6; 62.1 | 44.7; 90.2 | 60.1; 91.6 |  | 16.6; 62.3 |  |  |  |
| **mVNT_50_** | GMT | -- | 58.6 | 228.2*** | 315.3*** |  | 41.8 |  |  |  |
|  | 95% CI | -- | 43.2; 79.3 | 161.4; 22.6 | 222.1; 447.5 |  | 27.7; 63.2 |  |  |  |
| **cVNT_50_** | GMT | 0 | 16.6 | 37.7*** | 42.8*** |  | 46.4 |  |  |  |
|  | 95% CI | 0 | 13.0; 21.2 | 26.7; 53.2 | 30.2; 60.8 |  | 31.5; 68.4 |  |  |  |

*AU/mL=anti-RBD IgG concentration expressed in arbitrary units/mL; Inhibition % RBD:hACE2 at a dilution 1/100;* *Molecular virus neutralization titre mVNT_50_, highest serum dilution for inhibiting 50% of RBD:HACE2 interaction; cVNT=conventional live-virus neutralization titre. Post-3^rd^ dose-Hom=third dose with SOBERANA 02. Post-3^rd^ dose-Het=third dose with SOBERANA Plus. GMT=Geometric Mean Titre. 25^th^-75^th^=25-75 percentile. CCSP=Cuban Convalescent Serum Panel; *p<0.05; *** p< 0.0005 (paired Student t test: mVNT_50_, cVNT; Wilcoxon signed rank test: anti-RBD IgG, % Inh RBD:hACE2) for comparison the response after third dose respect to second dose.*

**Table VI. Kinetic of immune response after two doses of SOBERANA 02 and a third homologous or heterologous dose (pooled analysis from Phase I and Phase IIa)**

|  | Immune response to first (days 14, 28) and second doses (days 42 and 56); baseline value: day 0 | | | | | Immune response to 3rd dose heterologous | | Immune response to 3rd dose homologous | | **CCSP** |
| --- | --- | --- | --- | --- | --- | --- | --- | --- | --- | --- |
| Days | 0 | 14 | 28 | 42 | 56 | 70 | 84 | 70 | 84 | **--** |
| N | 120 | 120 | 117 | 113 | 110 | 58 | 58 | 51 | 51 | **68** |
| Seroconversion (%) | -- | 16.7 | 21.4 | 84.1 | 76.1 | 98.3 | 98.3 | 96.1 | 98.0 | **--** |
| IgG (AU/mL) Median | 1.95 | 2.52 | 3.1 | 27.0 | 23.7 | 99.3 | 113.4 | 71.6 | 81.7 | **50.8** |
| 25^th^-75^th^ percentile | 1.95; 1.95 | 1.95; 6.8 | 1.95; 9.0 | 12.8; 94.9 | 9.5; 85.5 | 49.9; 295.3 | 66.3; 339.5 | 34.7; 181.4 | 37.7; 208.2 | **23.8; 94.0** |
| Wilcoxon signed rank test vs. T0 | -- | 1.31E-12 | 6.41E-14 | 8.75E-20 | 8.75E-20 | 5.15E-11 | 3.50E-11 | 5.14E-10 | 5.14E-10 | **--** |
| Wilcoxon signed rank test vs. T56 | -- | -- | -- | -- | -- | 3.19E-10 | 6.71E-11 | 1.88E-9 | 1.09E-9 | **--** |

*Vaccination schedule: three doses (on T0, T28, T56). N: number of participants’ sera evaluated*

*Seroconversion****:*** *4-fold increment of anti-RBD IgG concentration at timepoint respect to pre-vaccination.*

*Anti-IgG are expressed as AU/mL (median; 25th-75th percentile).*

**Table VII. Anti-RBD IgG antibodies inhibit the interaction between RBD and human ACE2 receptor after two doses of SOBERANA 02 and a third homologous or heterologous dose (pooled analysis from phase I and phase IIa).**

|  | Immune response to first (days 14, 28) and second doses (days 42 and 56);  baseline value: day 0 | | | | Immune response to 3rd dose homologous | | | Immune response to 3rd dose heterologous | | | **CCSP** | |
| --- | --- | --- | --- | --- | --- | --- | --- | --- | --- | --- | --- | --- |
| Days | 0 | 14 | 42 | 56 | | 70 | 84 | | 70 | 84 | |  |
| N | 119 | 116 | 106 | 112 | | 58 | 58 | | 51 | 51 | | **68** |
| Inhibition (%) Median | 4.7 | 6.9 | 33.3 | 29.1 | | 82.1 | 79.8 | | 72.1 | 69.9 | | **32.0** |
| 25^th^-75^th^ percentile | 1.3; 7.8 | 3.4; 10.3 | 13.2; 70.0 | 12.7, 60.3 | | 65.2; 92.5 | 60.9; 92.0 | | 44.4; 89.0 | 42.6; 90-1 | | **16.6; 62.3** |
| Wilcoxon signed rank test vs. T0 | -- | 2.88E-7 | 7.79E-18 | 7.54E-19 | | 3.69E-11 | 3.69E-11 | | 7.56E-10 | 7.56E-10 | | **--** |
| Wilcoxon signed rank test vs. T56 | -- | -- | -- | -- | | 6.032E-11 | 9.372E-11 | | 7.552E-10 | 1.182E-9 | | **--** |
| mVNT_50 (_GMT) | 2 | 8.9 | 57.9 | 56.0 | | 345.1 | 319.0 | | 220.2 | 202.2 | | **41.8** |
| CI 95% | 2 ;2 | 7.3;10.8 | 42.6; 78.8 | 42.4; 74.0 | | 234.9; 507.1 | 231.5;439.6 | | 155.6; 311.6 | 141.9; 288.0 | | **27.7; 63.2** |
| paired Student t test (vs. T0) | 1.33E-28 | 1.21E-40 | 2.75E-45 | 2.16E-63 | | 7.10E-38 | 3.12E-31 | | 7.07E-31 | -- | |  |
| paired Student t test (vs. T56) | -- | -- | -- | 1.32E-35 | | 2.61E-23 | 1.21E-14 | | 1.94E-14 | -- | |  |

*Vaccination schedule: three doses (on T0, T28, T56). N: number of participants’ sera evaluated*

*% of inhibition of RBD: hACE2 interaction at 1/100 serum dilution (median, 25^th^-75^th^ percentile). Molecular virus neutralization titre mVNT_50_, highest serum dilution for inhibiting 50% of RBD:HACE2 interaction; (GMT, IC 95%). CCSP: Cuban Convalescent Serum Panel;*

**Table VIII. Neutralizing titre of SARS-CoV-2 live-virus after two doses of SOBERANA 02 and a third homologous or heterologous dose (pooled analysis from phase I and phase IIa).**

| Neutralizing titre cVNT50 at baseline (T0)  and after second dose (T56) | | | Neutralizing titre cVNT50 after third dose (T84) | | |
| --- | --- | --- | --- | --- | --- |
|  |  |  | 3th dose heterologous | 3th dose homologous | **CCSP** |
| Days | 0 | 56 | 84 | 84 |  |
| N | 106 | 89 | 55 | 51 | **68** |
| GMT | 2 | 12.5 | 42.5 | 32.8 | **46.4** |
| CI 95% GMT | 2 ; 2 | 9.6; 16.1 | 30.4; 59.4 | 23.8; 45.3 | **31.5; 68.4** |
| paired Student t test (vs. T0) | -- | 1.1868E-24 | 2.8029E-22 | 9.2253E-22 | **--** |
| paired Student t test (vs. T56) | -- | -- | 1.3695E-14 | 1.739E-14 | **--** |

*Conventional live-virus neutralization titre cVNT50; (GMT, IC 95%). N: number of participants’ sera evaluated*

*CCSP: Cuban Convalescent Serum Panel*.

**Table IX. Comparison of immune response induced by age groups after three doses (only Phase IIa)**

| Age group | Anti-RBD IgG Concentration | Inhibition (%) of RBD:hACE2 interaction | Molecular neutralization  (mVNT_50_) | Neutralization  viral  (cVNT_50_) |
| --- | --- | --- | --- | --- |
|  | Median  (25^th^-75^th^ Percentile) | Median  (25^th^-75^th^ Percentile) | GMT  (CI 95%) | GMT  (CI 95%) |
| 19-59 y/o  N=72 | 98.7  (58.5; 238.8) | 79.7  (57.0; 91.4) | 309.3  (235.0; 406.9) | 44.9  (33.0; 61.2) |
| 60-80 y/o  N=24 | 71.6  (27.9; 156.6) | 74.1  (39.9; 83.8) | 166.1  (98.3; 280.6) | 30.5  (20.2; 46.0) |
| Mann-Whitney U | 0.063 | 0.099 | 0.037* | 0.126 |

** Student t test N: number of participants’ sera evaluated.*

**Table X. Bivariate correlation after three doses between immune variables (pooled analysis from Phase I and Phase IIa)**

|  | | | anti-RBD IgG | mVNT_50_ | % RBD:hACE2 Inhibition |
| --- | --- | --- | --- | --- | --- |
| 3th dose Heterologous | IgG anti-RBD | Spearman r^2^ | 1.000 | 0.927^**^ | 0.919^**^ |
|  |  | Sig. (2-tailed) | . | 0.000 | 0.000 |
|  |  | N | 58 | 58 | 58 |
|  | mVNT_50_ | Spearman r^2^ | 0.927^**^ | 1.000 | 0.978^**^ |
|  |  | Sig. (2-tailed) | 0.000 | . | 0.000 |
|  |  | N | 58 | 58 | 58 |
|  | % RBD:hACE2 Inhibition | Spearman r^2^ | 0.919^**^ | 0.978^**^ | 1.000 |
|  |  | Sig. (2-tailed) | 0.000 | 0.000 | . |
|  |  | N | 58 | 58 | 58 |
|  | cVNT_50_ | Spearman r^2^ | 0.888^**^ | 0.922^**^ | 0.903^**^ |
|  |  | Sig. (2-tailed) | 0.000 | 0.000 | 0.000 |
|  |  | N | 55 | 55 | 55 |
| 3th dose homologous | IgG anti-RBD | Spearman r^2^ | 1.000 | 0.817^**^ | 0.800^**^ |
|  |  | Sig. (2-tailed) | . | .000 | .000 |
|  |  | N | 51 | 51 | 51 |
|  | mVNT_50_ | Spearman r^2^ | 0.817^**^ | 1.000 | 0.992^**^ |
|  |  | Sig. (2-tailed) | 0.000 | . | 0.000 |
|  |  | N | 51 | 51 | 51 |
|  | % RBD:hACE2 Inhibition | Spearman r^2^ | 0.800^**^ | 0.992^**^ | 1.000 |
|  |  | Sig. (2-tailed) | 0.000 | 0.000 | . |
|  |  | N | 51 | 51 | 51 |
|  | cVNT_50_ | Spearman r^2^ | 0.749^**^ | 0.928^**^ | 0.936^**^ |
|  |  | Sig. (2-tailed) | 0.000 | 0.000 | 0.000 |
|  |  | N | 51 | 51 | 51 |

*** Correlation is significant at the 0.01 level (2-tailed); r^2^: Correlation Coefficient*

| **Figure I. Risk-Benefit Balance.**  *Benefit_1_ = Proportion of individuals with seroconversion (≥4-fold increase in antibody concentration over pre-immunization levels) at 84 days.*  *Risk_1_ = Grade-3 adverse events vaccine-related.* | |
| --- | --- |
| **3rd dose heterologous** | **3rd dose homologous** |
| 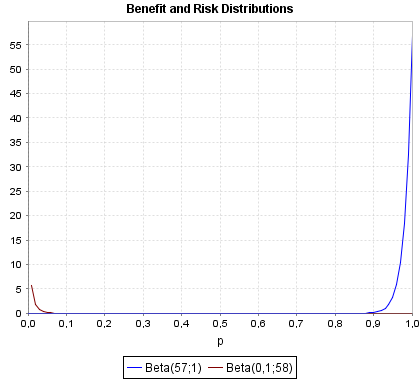  **Bayes Factor (likelihood ratio): 491.5** | 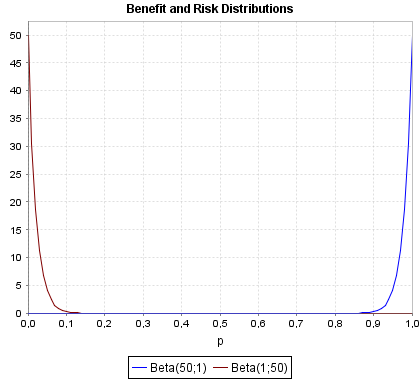  **Bayes Factor (likelihood ratio): 49.0** |
| *Benefit_2_ = Proportion of individuals with cVNT ≥ 20 at 84 days.*  *Risk_1_ = Grade-3 adverse events vaccine-related.* | |
| **3rd dose heterologous** | **3rd dose homologous** |
| 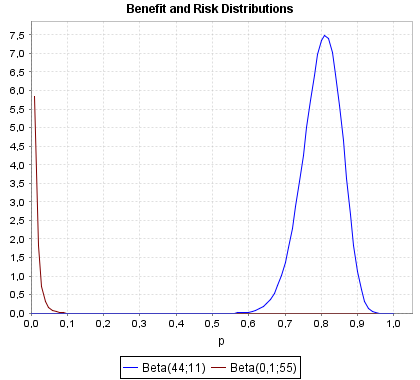  **Bayes Factor (likelihood ratio): 400.0** | 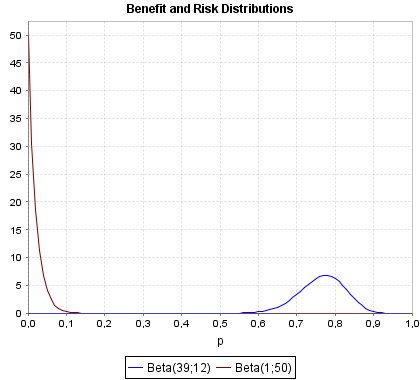  **Bayes Factor (likelihood ratio): 38.2** |

***─*** *Benefit Distributions (blue line);* ***─*** *Risk Distributions (red line)*

**Appendix C. Immunological Techniques**

*C.1 Anti-RBD IgG response:* Anti-RBD IgG in sera was evaluated by a quantitative ultramicro ELISA (UMELISA SARS-CoV-2 anti- RBD, Centre for Immunoassay, Havana, Cuba) using d-RBD as coating antigen (4 µg/mL) and an in-house standard characterized serum, which was arbitrarily assigned 200 AU/mL (based on a half-maximal inhibitory titre of 200 and a conventional virus neutralization titre of 160). The standard curve comprised six two-fold serial dilutions (0, 4, 8, 16, 32 and 64 AU/mL) of the standard. Samples were evaluated in duplicate. After incubation step, two separated steps of biotin-conjugate anti-IgG human (0.1 µg/mL) (Sigma Aldrich, San Luis, EE UU) and later streptavidin/alkaline-phosphatase (Roche, Basel, Swiss) in appropriate buffer were added. The final fluorimetric reaction was induced by adding substrate 4-Methylumbelliferyl Phosphate (Sigma Aldrich, San Luis, EE UU). The reference curve was constructed using a linear interpolation function. The concentration of anti-RBD IgG was expressed as AU/mL. The seroconversion rate was calculated by dividing the concentration at each time point (at Tx) by the pre-vaccination concentration (at T0). A rate ≥ 4 was considered as seroconversion.

*C.2 Molecular virus neutralization test*: Microtitre plates were coated with 250ng/well of ACE2-hFc in carbonate-bicarbonate buffer, 0.1M (pH9.6) and incubated overnight at 4°C. Plates were blocked with 200µL/well of 2% of skim milk in PBST (PBS with Tween 20 0.05%) during 1 h, at 37°C. Serial dilutions of sera were pre-incubated with RBD-mouse-Fc (RBD-mFc) at a final concentration of 20 ng/mL, for 1 h at 37^o^C. Mixtures were added to the plates and incubated for 2 h at 37°C. The binding of RBD-mFc was detected by addition of alkaline phosphatase (AP) conjugated anti-mouse IgG antibody (Sigma) for 1 h at 37°C. Finally, p-nitrophenylphosphate (Sigma) at 1 mg/mL in diethanolamine buffer (pH9.8) was added, and plates were incubated at RT for 30 min. The OD at 405nm was measured using a microwell system reader (BioTek). In all steps other than blockade, samples and reagents were added using a final volume of 50 µL/well. Three washing steps with PBST followed all incubations. RBD-mFc, sera and antibody conjugates were diluted in skim milk 0.2%/PBST. Inhibition was calculated and expressed as percentage according to the next formula: Inhibition (%)= [1-(OD_405nm_sample/OD_405nm_ maximal recognition)] x 100. Herein, maximal recognition corresponds to wells incubated only with RBD-mFc (20 ng/mL). For determination of the mVNT_50_, dilutions were log transformed and the highest dilution giving 50% of inhibition was calculated.

*C.3. Phages construction*

RBD-coding sequence (Arg328-Thr533) was synthesized by Eurofins^TM^ (Germany) and cloned in the phagemid vector pCSM through ApaLI and NotI restriction sites. The original gene was modified by site-directed mutagenesis to introduce the triplets coding for the desired replacements in the RBD sequence. Phages displaying D614G-RBD and variant Delta-L452R+T478K were rescued with M13KO7 helper phage. Display levels were determined by enzyme-linked immunosorbent assay (ELISA) on microplates coated with 9E10 mAb, an antibody targeting the *c-myc* tag fused to the carboxyl-terminal end of displayed proteins. Phages displaying D614G RBD were used as reference, assuming a display level of 100 units/mL, whereas relative levels of mutated variants were subsequently calculated. Phage-displayed RBDs were biologically active by ELISA on microplates coated with the recombinant extracellular region of human ACE-2 receptor fused to human Fc (rACE-2/Fc), confirming their proper folding on viral particles assembled in *E.coli* periplasm.

*C.4. Molecular Virus Neutralization Test with phage-displayed RBDs*

The inhibition of RBD/ACE2 interaction by anti-RBD antibodies in sera from 16 vaccinated individuals (first and second doses: 25 µg SOBERANA 02, third dose: SOBERANA Plus heterologous scheme) was evaluated in polyvinyl chloride microplates coated with rACE-2/Fc at 5 μg/mL in phosphate-buffered saline (PBS) overnight at 4ºC and blocked for 1h with PBS containing 4% skim milk powder (M-PBS) at room temperature. Each phage preparation was diluted to 0.075 display units/mL in M-PBS containing serially 4-fold diluted sera (from 1/25 to 1/25 600). Phages diluted similarly in M-PBS alone were used as controls for maximum binding (100%). After 1h pre-incubation at RT, diluted phages were incubated on coated/blocked microplates during an additional hour at RT. Plates were washed with PBS containing 0.1% Tween 20 (PBS-T) and incubated 1h at RT with an anti-M13 mAb conjugated to horseradish peroxidase (GE Healthcare, USA), properly diluted in M-PBS. After washing, the substrate solution (500 μg/ml ortho-phenylenediamine and 0.015% hydrogen peroxide in 0.1 mol/l citrate-phosphate buffer, pH 5.0) were added. The reaction was stopped 15 min later, with 2.5 mol/l sulfuric acid. Absorbances (Abs) at 490 nm were determined with a microplate reader. Relative binding levels were calculated as the ratio between Abs at 490 nm and the maximum binding signal of the same phage-displayed RBD variant in M-PBS. Sigmoidal inhibition curves were analyzed with GraphPad Prism 5 software, and serum dilutions producing 50% binding inhibition (mVNT_50_) were calculated for each serum sample against each phage-displayed RBD variant.

*C.5 Conventional Virus Neutralization test*

Neutralizing antibodies against live SARS-CoV-2 was performed in a biosecurity laboratory level 3 (National Civil Defense Research Laboratory, Havana, Cuba) by the conventional virus neutralization test, the gold standard for determining antibody efficacy against SARS-CoV-2. Serial dilutions of heat-inactivated serum samples (starting from 1:5) in Eagle’s Minimal Essential Medium (Gibco, UK) containing 2 % fetal bovine serum (Capricorn, Germany) were incubated for 1 hour at 37°C with an equal volume of virus solution containing 100 TCID_50_ of SARS-CoV-2 (Strain 2025, variant D614G, Cuban Collection at National Civil Defense Research Laboratory) in cell plates containing a semiconfluent VeroE6 monolayer (10^4^ cell/well). The highest serum dilution, showing an OD at 540 nm representing the 50% of average OD values from control cell wells (VeroE6 monolayer with mixture of virus-serum) was considered as the neutralization titre and is represented as neutralizing titre 50 (cVNT50).

*C.6 Specific T-cell response*

Peripheral blood mononuclear cells (PBMCs) were isolated from a subset (n=24) of vaccinated subjects, at on day 0 (before first immunization); day 56 (28 days after the second dose) and 84 (28 days after the third dose), homologous scheme: n=13, heterologous scheme: n=11). RBD-specific T-cell response producing IFN- γ and IL-4 were quantified with enzyme-linked immunospot (ELISpot) assay using human IFN-γ ELISpot^PLUS^ HRP kit (Mabtech, Sweden) and human IL-4 ELISpot^plus^ HRP kit (Mabtech, Sweden) following the manufacturer´s instructions. Briefly, cryopreserved peripheral blood mononuclear cells from vaccinated subjects were thawed and rested for 6 hours in RPMI 1640 medium supplemented with 1000 units/mL penicillin, 1 mg/mL streptomycin, 1 mM pyruvate (Gibco), 50 µM β-mercaptoethanol (Sigma-Aldrich) and 10% *(v/v)* heat-inactivated fetal calf serum (Capricorn). PBMCs (2.5 x 10^5^ cells/well, in duplicate) were stimulated with recombinant RBD (10 µg/mL) at 37°C for 24 h before detection. Specific T-cell response was expressed as the number of spot-forming cells per 10^6^ cells; measurements were subtracted from the unstimulated control values.
